# Part 2 of a 2-part open-label clinical trial evaluating repeated nicotinamide riboside parenteral administration: 90 days of at-home subcutaneous injections

**DOI:** 10.64898/2026.10.13.26365470

**Authors:** Anne Russ, Yasmeen Nkrumah-Elie, Frehiwet Hailu, Jun Kwon, Tracy Allen, Andrew Shao, Halland Chen

## Abstract

Nicotinamide riboside (NR), a precursor to the essential coenzyme nicotinamide adenine dinucleotide (NAD+), has been extensively studied following oral administration, while the longer-term safety and physiological effects of injectable NR remain less characterized. Following an initial in-clinic phase evaluating repeat intramuscular (IM) and subcutaneous (SC) administration of pharmaceutical-grade NR, participants transitioned to an at-home phase designed to assess the safety, feasibility, and exploratory physiological effects of repeated self-administered SC administration. Participants continued their assigned 50 or 100 mg dose group and self-administered NR subcutaneously 3 times weekly for approximately 90 days. Safety was assessed through adverse event (AE) monitoring, vital signs, and clinical laboratory health measures. Exploratory assessments included markers of mitochondrial bioenergetics, urinary biomarkers of oxidative stress, and participant-reported outcomes related to fatigue, sleep, pain, and quality of life. At-home administration of NR was generally well-tolerated, with no clear dose-dependent safety concerns identified over the study period. Further, participants were able to successfully self-administer repeated SC injections over the study period, demonstrating positive feasibility. Whole-blood NAD+ concentrations remained largely stable over time in both dose groups. MeScreen analysis demonstrated a reduction in GlycoATP at Day 30 and increases in the MitoATP/GlycoATP ratio at Day 30 and Day 90, signifying a preferential shift toward oxidative metabolism. These findings may be indicative of positive metabolic health shifts. Urinary oxidative stress profiling demonstrated heterogeneous temporal responses. Several lipid oxidation markers significantly declined at Day 30 but returned toward baseline at Day 90. Select markers of nucleic acid, nitrative stress, and advanced glycation end-product oxidation demonstrated more sustained reductions. These results for which the majority of measured oxidative stress markers improved suggest that NR injections may exhibit anti-oxidative characteristics. Validated participant-reported sleep and fatigue measures showed improvements. Self-reported facial skin improvement increased descriptively from 11.4% at baseline to 27.8% at Day 30 and 36.8% at Day 90; improvements noticed by others increased from 9.1% to 23.7%, while reported body-skin improvement increased more modestly, from 8.8% to 16.7%. As these outcomes were not designed or powered to establish efficacy or definitive clinical claims, the findings should be considered hypothesis generating and provide preliminary data to inform the selection of outcomes and design considerations for future randomized, placebo-controlled, adequately powered trials of SC NR administration.

**Significance Statement:** This is one of the first clinical trials evaluating safety and exploring the efficacy of nicotinamide riboside (NR) injections over an extended period. A favorable safety profile was observed for self-administered injections at doses of 50 and 100 mg. Quantitative changes in markers of mitochondrial efficiency and oxidative stress, and perceived self-reported improvements in functional outcomes of fatigue, sleep, and quality-of-life were independent of whole blood NAD+ measurements. This study is hypothesis generating and requires validation in future clinical trials.

## 1. Introduction

Nicotinamide adenine dinucleotide (NAD+) is an essential coenzyme that occupies a central role in cellular metabolism. Beyond its canonical role as a redox coenzyme in cellular energy production, NAD+ is also a required substrate for protein families involved in DNA damage repair, innate immune responses, and post-translational protein modification^1^. A growing body of evidence has established that aging is accompanied by progressive reductions in cellular and tissue NAD+ concentrations, although this relationship may be tissue and compartment dependent^2–7^. Indeed, recent human studies have challenged the notion of a universal age-related decline in whole blood NAD+ concentrations^8^. Nevertheless, in human observational studies, lower blood and skeletal muscle NAD+ levels are related to impaired physical performance, poor muscle health, frailty, and reduced mitochondrial content^9,10^. NAD+ homeostasis is further dysregulated in the presence of metabolic stressors, including overnutrition, excess alcohol intake, environmental contaminants, sedentary lifestyles, and various disorders^2,3,11–14^. The downstream physiological implications of diminished NAD+ availability are broad, having been linked to neurodegenerative conditions, musculoskeletal dysfunction, metabolic disorders, and inflammatory diseases^5,13,15^. From a molecular mechanistic perspective, converging lines of evidence have identified NAD+ as a central mediator of the hallmarks of aging, including genomic instability, mitochondrial dysfunction, chronic inflammation, and loss of proteostasis^16^. Additionally, diminished NAD+ levels are implicated in oxidative stress, a driver of cellular energy decline and damage, that has been linked to physiological manifestations of aging^17–19^. The restoration of NAD+ levels has therefore emerged as a potential therapeutic target,^4^ with the expectation that it may counteract the deleterious consequences associated with its decline^5^.

Nicotinamide riboside (NR) supplementation is a promising strategy to augment intracellular NAD+ levels^20,21^, with the potential to mitigate NAD+ age-related decline and associated conditions. NR is a naturally occurring pyridine-nucleoside B3 vitamin that enters cells and is directed toward the salvage pathway, where it is converted to NAD+ through a two-step process involving the enzyme of nicotinamide riboside kinase (NRK)^22^. Oral NR supplementation has repeatedly demonstrated safety and NAD+ elevation in multiple animal and human studies^23–25^. Typically, 10–14 days of daily oral NR intake are required to reach steady state NAD+ levels in whole blood, likely indicative of systemic equilibrium^25^. Of note, a meaningful fraction of orally administered NR may undergo gastrointestinal conversion mediated by BST1 (bone marrow stromal cell antigen 1 or CD157) and microbiota-driven biotransformation to nicotinamide and nicotinic acid prior to reaching systemic circulation, which introduce variability in systemic exposure ^26–28^. These characteristics of oral delivery raise the possibility that parenteral administration, which bypasses gastrointestinal metabolism, may offer advantages under circumstances where an immediate or consistent systemic boost to NAD+ is desirable. The safety and NAD+-raising effects of parenteral NR, specifically subcutaneous (SC) and intramuscular (IM) administration, remain poorly characterized. Until recently, pharmaceutical-grade NR (Niagen® Plus) appropriate for injectable research was unavailable. As a result, clinical investigation of alternative delivery routes has been limited.

To date, the available peer-reviewed human evidence on parenteral NR has been limited to intravenous (IV) administration. The first controlled human data on parenteral NR were reported in a preprint by Hawkins et al., who demonstrated that IV NR was generally well tolerated, without untoward effects on biochemical parameters^29^. Reyna et al^30^ later corroborated these findings in a retrospective clinical study of commercial wellness clients. In addition, preclinical investigations in a preprint report by Kwon et al.^31^ identified differential toxicologic profiles across IV, SC, and IM routes of NR administration, providing early evidence that the response to NR may vary as a function of delivery route. Relative to intravenous delivery, SC and IM administration each differ in their absorption profiles, with the latter routes generally characterized by more gradual uptake into circulation rather than direct vascular delivery, though generally more rapid and predictable than oral administration ^32^. In addition to potential differences in kinetics, SC and IM injections are likely more accessible alternatives to IV infusions and more suitable for outpatient, independent use. To date, however, no peer-reviewed human study has explored the safety, tolerability, or physiological effects of SC or IM NR administration. Therefore, the purpose of the present study was to address this gap by evaluating the safety and whole-blood NAD+ augmentation potential of repeated NR injections, stratified by dose and route of administration.

The parent investigation was designed as a two-part, open-label pilot study evaluating pharmaceutical-grade NR chloride (Niagen Plus). The present report focuses on the subsequent at-home phase (‘Part 2’) of the study. During the initial in-clinic phase (‘Part 1’), participants received three consecutive clinician-administered injections of 50 or 100 mg NR by either IM or SC route, followed by a 7-day washout. These findings established short-term safety under clinical supervision. However, it did not establish safety under conditions in which injectable NR would more likely be used, namely, repeated, self-administered dosing over weeks to months. Therefore, to address this open question, all participants then transitioned to self-administered SC injections three times weekly for approximately 90 days, at their originally assigned 50- or 100-mg NR dose, and were assessed at various timepoints.

The primary objective of Part 2 was to characterize the safety and tolerability of repeated at-home SC administration of 50 and 100 mg NR. Secondary and exploratory objectives were to evaluate the longitudinal changes in biomarkers of mitochondrial bioenergetics and oxidative stress, as well as participant-reported measures of fatigue, sleep, quality of life, and perceived skin changes during extended NR administration. These exploratory endpoints were included based on the known role that NAD+ has in mitochondrial homeostasis, antioxidant defense, and genomic maintenance, and its potential clinical applications on broader health and wellness outcomes ^1,5,7,33,34^. For exploratory outcomes not assessed at Part 2 Day 1, the original study baseline at Visit 1 was retained as the reference timepoint. Given the pilot design and modest sample size, these results, in addition to dose-stratified and sex-stratified subanalyses, were intended to generate hypotheses for future adequately powered, placebo-controlled studies, rather than definitively establish treatment efficacy or sex-specific effects of SC NR.

## 2. Materials and Methods

### 2.1 Trial Design

The parent investigation was a single-site, prospective, randomized, open-label pilot study evaluating the safety and biological effects of Niagen® Plus (ChromaDex, Inc., a Niagen Bioscience company, Los Angeles, California, USA), a pharmaceutical-grade form of nicotinamide riboside chloride compounded as a sterile powder for injection by Wells Pharmacy Network (Houston, TX, USA).

The study consisted of two sequential phases. In Part 1, participants were randomized to receive either 50 or 100 mg NR by IM or SC injections on three consecutive study days (Visits 1-3), followed by a 7-day washout, as described in Nkrumah-Elie, 2026.

Participants returned for Visit 4 (study Day 10), which served as Day 1 of Part 2. At this visit, they underwent clinical assessments and received training and instructions in SC self-administration. All participants continued at their originally randomized dose, however, all transitioned to the SC route of administration, self-administered three times weekly at home for 90 days. Participants returned to the clinic at Part 2 Day 30 (V5) and Part 2 Day 90 (V6, and last day of administration) for clinical assessments, and received a follow-up call on approximately 14 days post-study. Throughout the present manuscript, Part 2 Day 1, Day 30, and Day 90 correspond to Visits 4, 5, and 6, respectively, or overall study Days 10, 40, and 100.

### 2.2 Study Participants

Participant recruitment procedures, inclusion and exclusion criteria, were described previously in Part 1 authored by Nkrumah-Elie, 2026. Briefly, participants were recruited through IRB-approved informational documents, community-based outreach, and public boards. Eligible participants were comprised of generally healthy adults aged 18 years or older with elevated fatigue at screening, defined as a Fatigue Assessment Scale (FAS) score of >22. Participants were also required to be naïve to NAD+ or NAD+ precursor injections and non-anemic. Key exclusion criteria included one or more uncontrolled chronic illness including but not limited to diabetes, cardiovascular, liver, or kidney disease, or any form of cancer. Additional exclusion criteria were as follows: an acute illness within 14 days prior to the first clinic visit, more than one chronic disease diagnosis under active treatment, a cancer diagnosis within the last 5 years, current pregnancy or lactation, the use of NAD+-related supplements within the 60 days prior to enrollment, a hypersensitivity or allergy to bacteriostatic water, NR, niacin, and other forms of vitamin B3/NAD+ precursors, excessive daily use of alcohol (defined as 4 or more drinks on one occasion), illicit drug use, and any other condition rendering the participant unsuitable per the Principal Investigator

Participants were randomized to 50 mg IM, 50 mg SC, 100 mg IM, or 100 mg SC, following consent, in a 1:1:1:1 ratio using a computer-generated allocation sequence without stratification; age and sex were not incorporated into the allocation process. Given the exploratory, pilot nature of the study and limited sample size, randomization without stratification was considered appropriate, with demographic balance assessed descriptively across groups. To facilitate real-time safety monitoring, the study was conducted in an open-label design, with both participants and clinicians unblinded to treatment assignments.

Participants agreed to maintain their typical diet and physical activity from two days prior to Visit 1 through completion of Visit 6. Prior to clinic visits, participants were instructed to consume ≥2 L of water, abstain from alcohol for 24 hours, and arrive fasted (minimum 8 hours, apart from water). Participant compensation was not provided, however, the reimbursement of transportation expenses to the clinic was offered.

This clinical investigation was conducted in accordance with the International Conference on Harmonisation Good Clinical Practice (ICH GCP), the Declaration of Helsinki, and the United States Code of Federal Regulations (21 CFR Parts 50 and 56, and 45 CFR 46). The study received full authorization from the Institute of Regenerative and Cellular Medicine’s Institutional Review Board (HHS/OHRP IRB00009500), Protocol number: NB-2025-001, Approval number: IRCM-2025-451. The trial was registered on ClinicalTrials.gov (NCT07251608). All participants provided written informed consent. An independent medical monitor (IMM) was contracted by the sponsor to serve as the primary safety monitor, assess the attribution of adverse events, and ensure the trial’s scientific integrity and regulatory compliance. The trial was administered by Impact Health Medical, in their clinic in Aventura, Florida. Recruitment started in September 2025 and the last participant out, as defined as the last follow-up conducted, was in February 2026.

### 2.3 Interventions

#### 2.3.1. Investigational product

Pharmaceutical-grade NR chloride (Niagen® Plus, ChromaDex, Inc., a Niagen Bioscience company, Los Angeles, California, USA), compounded by Wells Pharmacy Network (Ocala, Florida, USA) was used for injection. The study material was provided as 500 mg sterilized powder in multi-use vials and was reconstituted with 5 ml or 10 ml of bacteriostatic water (vendor), per participant, by the on-site clinician. The reconstituted material was stored under refrigerated conditions between uses.

#### 2.3.2. At-Home Intervention

At Visit 4, study personnel trained participants on proper administration of the investigational product subcutaneously in the abdomen and upper thigh, in an alternating fashion, utilizing standard insulin syringes with 29-gauge needle. In addition, they provided instructions for reconstitution of the 500 mg of NR in multiuse vials with bacteriostatic water, as well as instructions to store the product under refrigerated conditions, and to only use 1 vial for up to 28 days.

Participants self-administered their assigned dose three times weekly for 90 days. Adherence was assessed through self-reporting from participants to the clinical staff and study coordinator.

### 2.4 Outcomes

#### 2.4.1 Primary Outcome Measures

The primary outcome of Part 2 was to evaluate the safety and tolerability of repeated self-administered subcutaneous NR at doses of 50 mg and 100 mg over 90 days. Safety outcomes included adverse events, injection-site reactions, vital signs, and clinical chemistry and hematology parameters.

Vital signs included systolic and diastolic blood pressure, heart rate, respiratory rate, peripheral oxygen saturation, and body temperature; each parameter was measured using calibrated devices with participants in a standardized seated position at Visits 4, 5, and 6. Venous blood and plasma were collected for comprehensive metabolic panel, complete blood count, homocysteine, and high-sensitivity C-reactive protein analyses during Visits 4, 5, and 6. Participant-reported tolerability, including injection-site pain and soreness, was also assessed throughout the study

#### 2.4.2 Secondary Outcome Measures

##### 2.4.2.2. Whole Blood NAD+ Pharmacokinetics

Quantitative capillary whole-blood NAD+ pharmacokinetics were assessed using dried blood spots (DBS; Revvity, Pittsburgh, PA). Clinicians collected DBS samples during clinic visits, and participants collected them during the washout period using clinic-provided materials and instructions. A lancet was applied to a clean finger, the first blood drop was gently wiped and at least one drop of blood was applied to a chemically coated dried blood spot card, halting enzymatic activity ^35,36^. The blood on the card was dried at room temperature for a minimum of 3 hours, away from direct sunlight, fans, or air vents. Once dry, the card was placed in a specimen bag with a desiccant, and placed in a refrigerator (for at-home collections) or in the -20°C freezer (for in-clinic collections). Samples were shipped to the vendor on dry ice, and analyzed using LC-MS/MS.

#### 2.4.3 Exploratory Outcome Measures

##### 2.4.3.1 Mitochondrial Function

To characterize mitochondrial health, MeScreen™ (Verséa Health, Inc, Tampa, FL, USA) was employed as a functional mitochondrial assessment platform to evaluate cellular bioenergetics via minimally invasive dried blood sampling. The assay provides a dynamic assessment of mitochondrial function by quantifying parameters associated with oxidative phosphorylation, glycolytic activity, and redox balance, incorporating measurements linked to oxygen consumption rate (OCR), extracellular acidification, and reactive oxygen species (ROS) production, which together provide a systems-level view of cellular energy metabolism.

MeScreen was assessed at Visit 1, Part 2 Day 30, and Part 2 Day 90.

##### 2.4.3.2. Urinary Oxidative Stress Markers

Oxidative stress was evaluated via a targeted panel of 16 urinary biomarkers spanning lipid peroxidation, DNA/RNA oxidation, protein oxidation, nitrative stress, and advanced glycation end-products (Vibrant, Santa Clara, CA, USA). Urine was self-collected by participants using a standard collection cup and subsequently pipetted into a storage tube. Urine was not restricted to first morning void and was stored at room temperature. Following extraction and purification, sample analysis was performed using liquid chromatography-tandem mass spectrometry (LC-MS/MS). The panel was optimized across three specific analytical sub-methods to accommodate the distinct chemical properties and ionization requirements of the respective analytes. The panel analyzed 11-β-prostaglandin F2α, 15(R)-prostaglandin F2α, 8-iso-prostaglandin F2α, glutathione 4-hydroxynonenal, malondialdehyde (MDA), 8-hydroxy-2-deoxyguanosine, 8-8-hydroxyguanine, 8-8-hydroxyguanosine, 8-nitroguanine, 8-nitroguanosine, 3-bromotyrosine, 3-chlorotyrosine, dityrosine, nitrotyrosine, Nε-carboxyethyllysine (CEL), Nε-(carboxymethyl)lysine (CML), and a composite oxidative stress score. Chromatographic separation and mass detection were conducted on either a Xevo TQ-XS or a Xevo TQ Absolute triple quadrupole mass spectrometer, each integrated with an ACQUITY UPLC system (Waters Corporation, Milford, MA, USA). All assays were executed within a CLIA-certified and CAP-accredited laboratory environment. Method precision was maintained with both intra- and inter-assay coefficients of variation (CV) of <20% for all monitored analytes. Reference intervals were established from a healthy adult cohort in strict accordance with Clinical and Laboratory Standards Institute (CLSI) guidelines, with data normalized to urinary creatinine, and expressed relative to creatinine.

Samples were collected at Visit 1, Part 2 Day 30, and Part 2 Day 90.

##### 2.4.3.5. Participant Reported Outcomes (PRO)

PROs pertaining to fatigue, sleep, and quality of life were assessed using validated PROMIS instruments listed in Table 1.

**Table 1.** Participant Reported Outcome measures – schedule and baseline definition. PROs were administered during the study as summarized below. The table defines the instrument, study timepoints, and baseline used for longitudinal analyses.

| **Instrument** | **Timepoints** | **Baseline used** |
| --- | --- | --- |
| PROMIS Fatigue Short Form 7b | Visit 1, Part 2 Day 30, Part 2 Day 90 | Visit 1 |
| PROMIS Sleep Disturbance-8b | Visit 1, Part 2 Day 30, Part 2 Day 90 | Visit 1 |
| PROMIS Sleep-Related Impairment-8a | Visit 1, Part 2 Day 30, Part 2 Day 90 | Visit 1 |
| PROMIS Global Health Scale V1.2 | Visit 1, Part 2 Day 30, Part 2 Day 90 | Visit 1 |
| FAS | Visit 1, Part 2 Day 30, Part 2 Day 90 | Visit 1 |

PROMIS instruments were scored per their respective manuals, and expressed as standardized T-scores, with a population mean of 50 and a standard deviation of 10. Higher T-scores indicate greater fatigue, sleep disturbance, and sleep-related impairment, whereas higher scores indicate better perceived quality of life for the Global Health Scale metrics, global mental health (GMH) and global physical health (GPH). The Fatigue Assessment Scale (FAS) was conducted during screening to support participant inclusion. A score of 22 indicated greater than average fatigue, warranting enrollment in the study.

An open-ended subjective skin questionnaire was administered at Visit 1, and Part 2 Days 1, 30, and 90. It addressed four domains: facial skin, facial skin noticed by others, body skin, and other aspects of physical appearance. Answers to open-ended questions were categorized as improvement, no change, worsening, other/neutral change, or unclear.

### 2.5 Statistical Analysis

Statistical analyses were performed using GraphPad Prism (GraphPad Software, LLC, Boston, MA, United States).

Part 2 Day 1 (Visit 4) served as the baseline for outcomes assessed at that visit. For outcomes not assessed at Visit 4 (MeScreen, urinary oxidative stress markers, and PROMIS), the study baseline (Visit 1) was retained. Changes in these outcomes therefore reflect cumulative exposure across Parts 1 and 2.

The primary analysis of longitudinal outcomes used a two-way repeated analysis of variance (ANOVA) test. Due to missed participant visits and unequal sample size between the groups, a mixed-model approach was utilized to accommodate incomplete measurements. When ANOVA testing resulted in statistical significance, post-hoc comparisons were performed using Tukey’s multiple comparison test to evaluate the statistical significance within- and between-groups. Statistical significance was defined as α = 0.05. For NAD+ metabolomics and exploratory outcome measures, sex- and dose-stratified analyses were also conducted.

## 3. Results

A total of 40 participants were randomized for the clinical trial (Figure 1). One participant was discontinued prior to the first injection due to a health concern identified by the PI, resulting in 39 participants in the intent-to-treat (ITT) population. The per-protocol population was defined as completion of all clinic visits and at least 80% of all assessments. All 39 participants met the criteria for ITT and they all completed the 6 clinical visits. However, two participants did not complete all assessments for all visits. (Figure 1). In addition, two other participants deviated from the assigned dosing during the at-home phase, with one increasing from 50 mg to 100 mg and the other reducing from 100 mg to 50 mg; therefore, these participants were analyzed in accordance with their actual doses.

**Figure 1.**
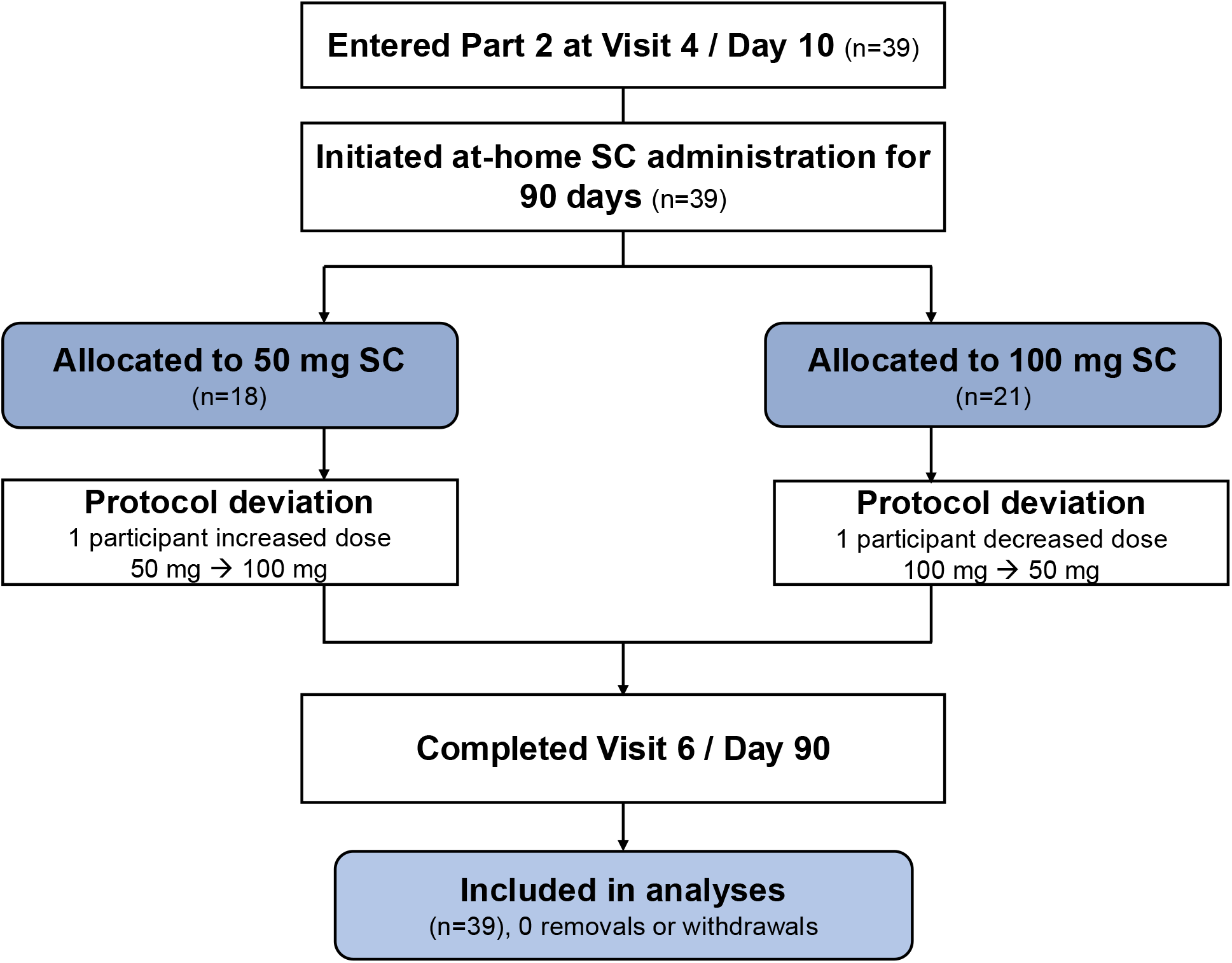
CONSOFT Flow Diagram - Part 2.

### 3.1 Demographics

**Table 2.**
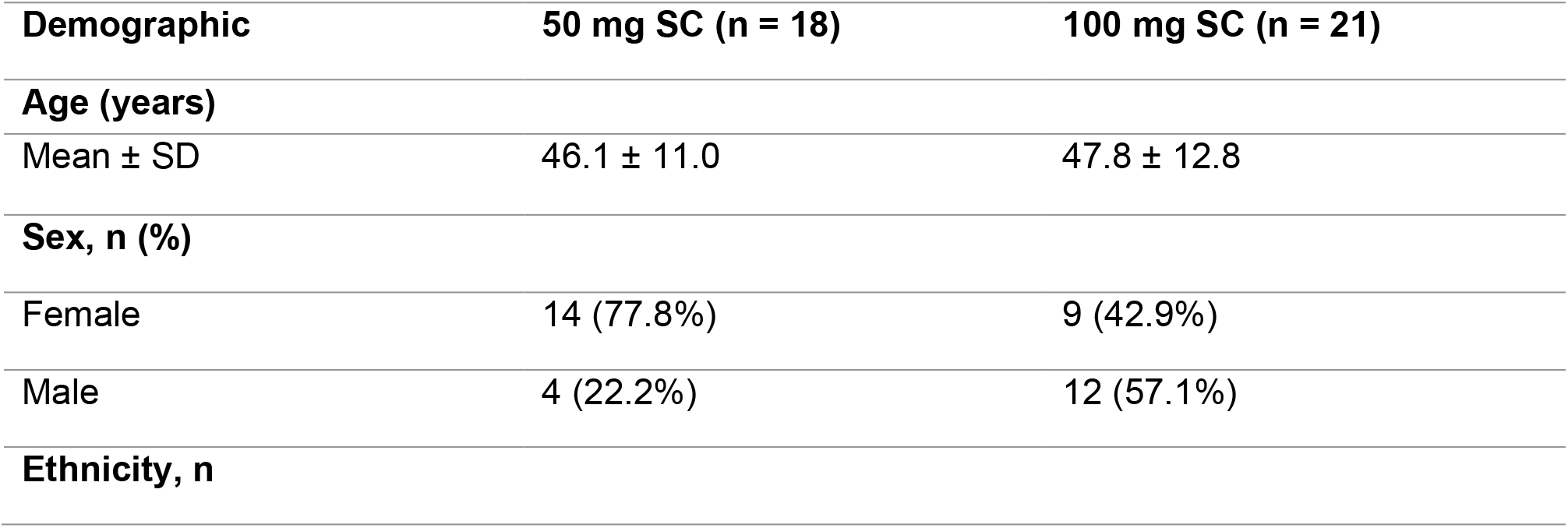
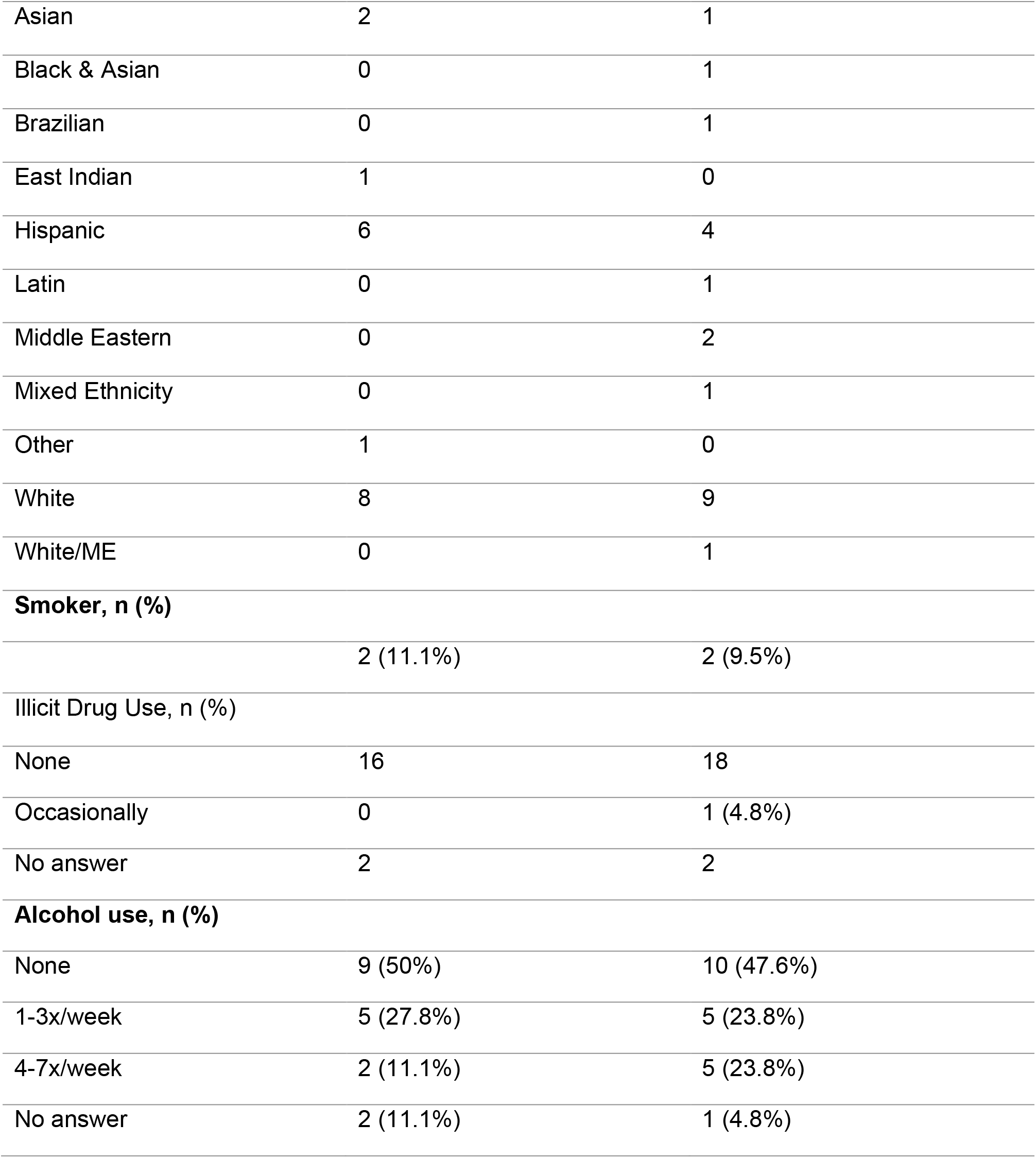
Participant demographics and baseline characteristics for Part 2.

Participant characteristics are presented according to the actual NR dose administered subcutaneously during the 90-day at-home phase. Participants who deviated from their original, assigned dose were analyzed in accordance with the dose they actually received. The Part 2 analysis population included 39 participants – 18 in the 50 mg SC group, and 21 in the 100 mg SC group.

### 3.2 Safety Assessments

#### 3.2.1 Vital Signs

Vital signs remained stable over the study period. Although several comparisons were statistically different, the observed changes were modest in magnitude, and no consistent pattern across vital parameters were observed (Table 3).

**Table 3.** Vital signs. Participants are grouped according to SC NR dose administered. BL represents Part 2 baseline assessment, followed by assessments at Day 30 and 90. Data are presented as mean ± SD. *p < 0.05 versus BL within the same dose group; †p < 0.05 versus the corresponding 50 mg group at the same timepoint. Note: only 37 participants were included in the analysis.

| Parameter | 50 mg SC |  |  | 100 mg SC |  |  |
| --- | --- | --- | --- | --- | --- | --- |
|  | BL | Day 30 | Day 90 | BL | Day 30 | Day 90 |
| Systolic blood pressure (mmHg) | 120.76 ± 15.03 | 127.00 ± 10.93 | 119.06 ± 17.49 | 124.10 ± 16.68 | 131.55 ± 12.09 | 134.00 ± 19.14† |
| Diastolic blood pressure (mmHg) | 75.59 ± 9.20 | 76.33 ± 6.99 | 76.35 ± 11.31 | 80.65 ± 15.42 | 81.30 ± 9.75 | 83.47 ± 14.34 |
| Heart rate (bpm) | 69.88 ± 10.79 | 70.67 ± 10.57 | 73.94 ± 9.85 | 74.45 ± 9.69 | 72.85 ± 10.74 | 73.05 ± 14.39 |
| Respiratory rate (breaths/min) | 16.12 ± 0.49 | 15.93 ± 1.22 | 15.53 ± 0.87* | 15.60 ± 0.82† | 16.10 ± 1.37 | 15.21 ± 3.51 |
| SpO<sub>2</sub> (%) | 98.47 ± 1.33 | 98.73 ± 0.96 | 98.65 ± 1.06 | 98.30 ± 1.49 | 98.75 ± 1.29 | 99.21 ± 0.98* |
| Temperature (°F) | 97.88 ± 0.52 | 97.59 ± 0.72 | 97.89 ± 0.48 | 97.99 ± 0.85 | 97.93 ± 0.71 | 97.59 ± 0.54 |

#### 3.2.2 Clinical Chemistry and Hematology

A total of 22 clinical chemistry and hematology markers were assessed throughout the study. Although statistically significant changes were observed for select measures, values generally remained within clinical reference ranges and were not considered clinically meaningful, or indicative of a safety signal. These findings were independently reviewed and confirmed by a third-party clinician.

Statistically significant changes were observed for select measures, MCHC, MCV chloride, carbon dioxide, calcium, and total bilirubin, across select timepoints. Between-group differences were also observed for hemoglobin and creatinine at baseline, creatinine and total bilirubin at Day 30, and hematocrit, creatinine, ALT, AST, and total bilirubin at Day 90 (p < 0.05). However, these changes were isolated and not consistently associated with dose or time-dependent patterns (Table 4).

**Table 4.**
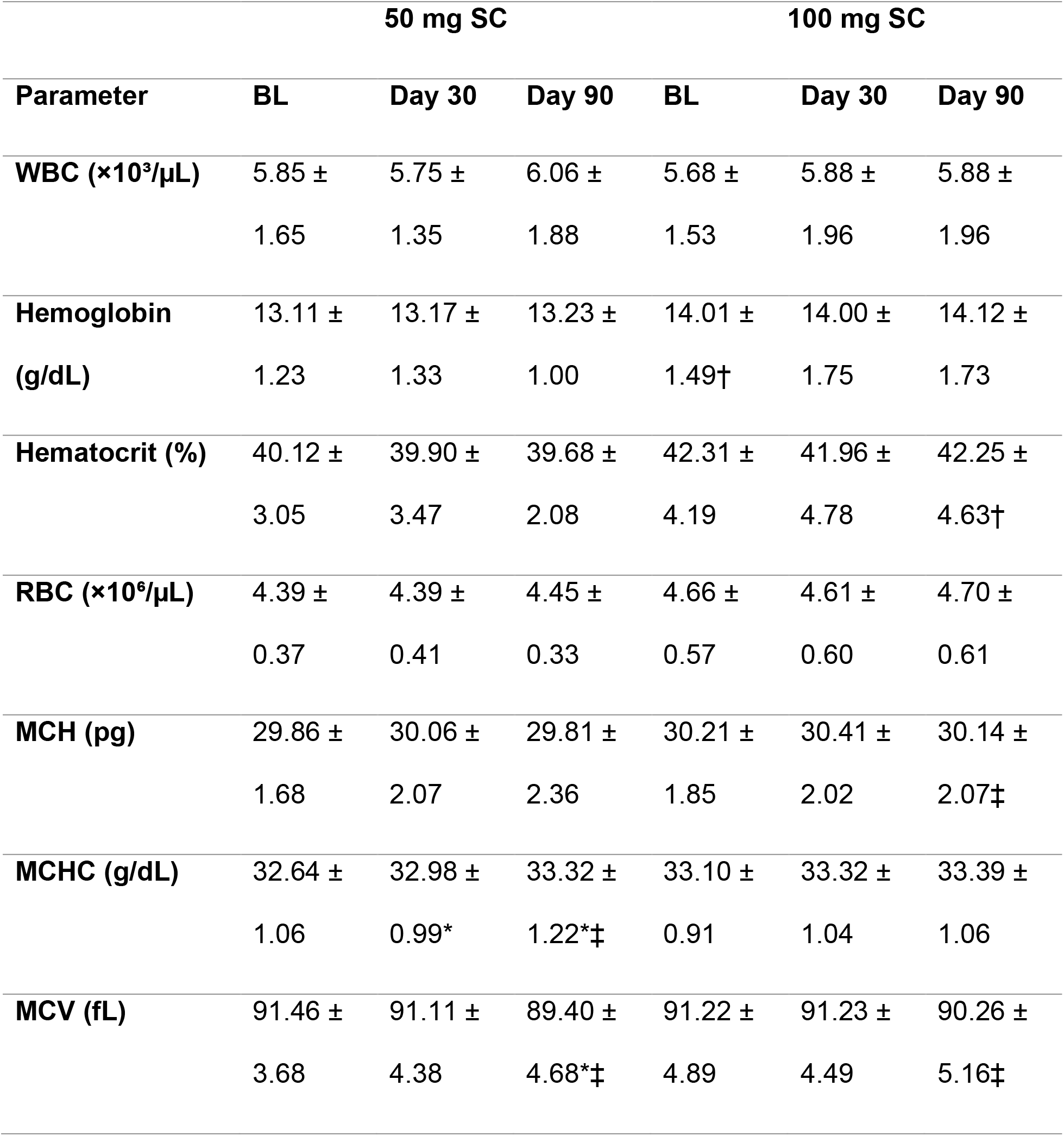
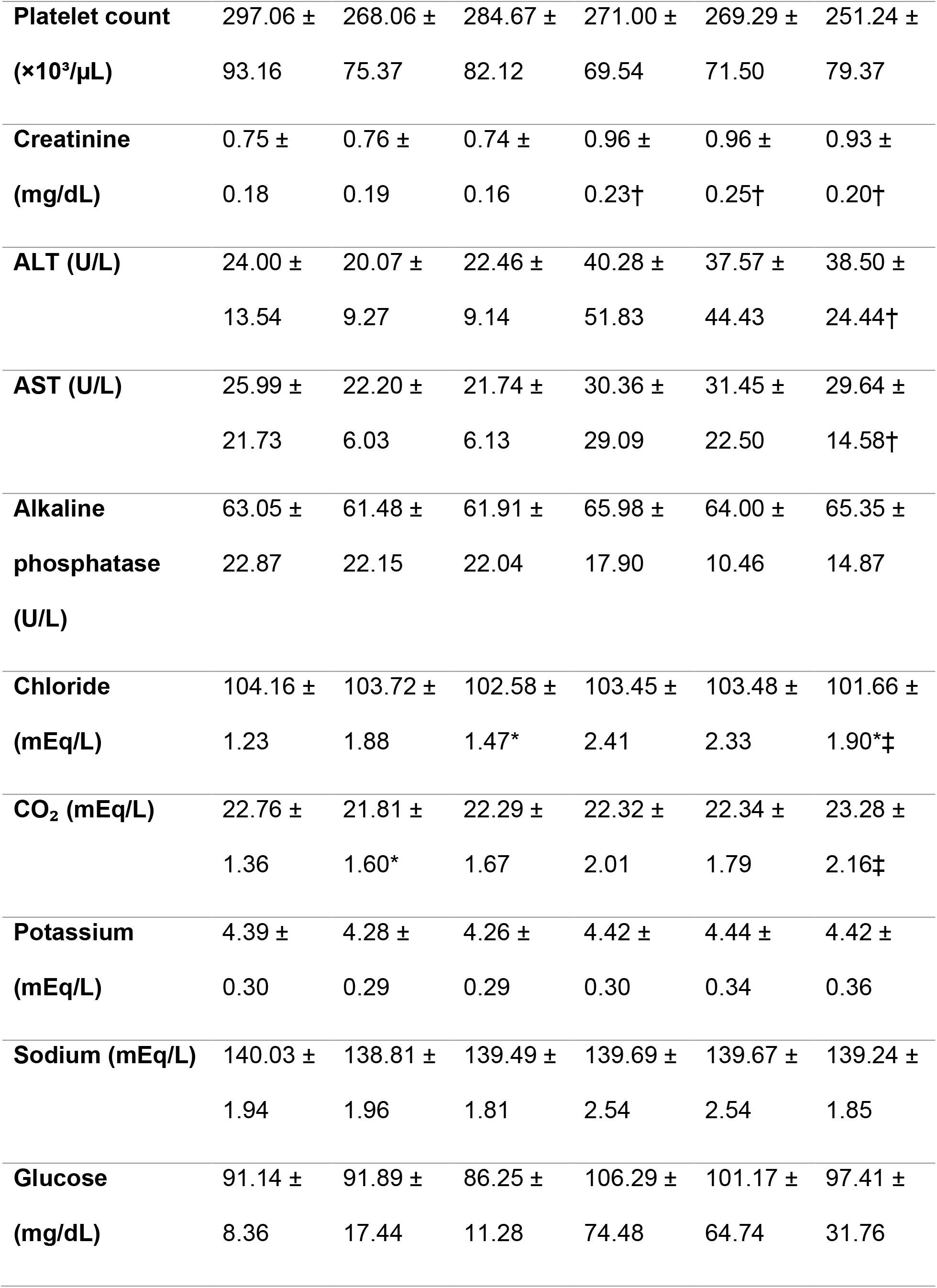
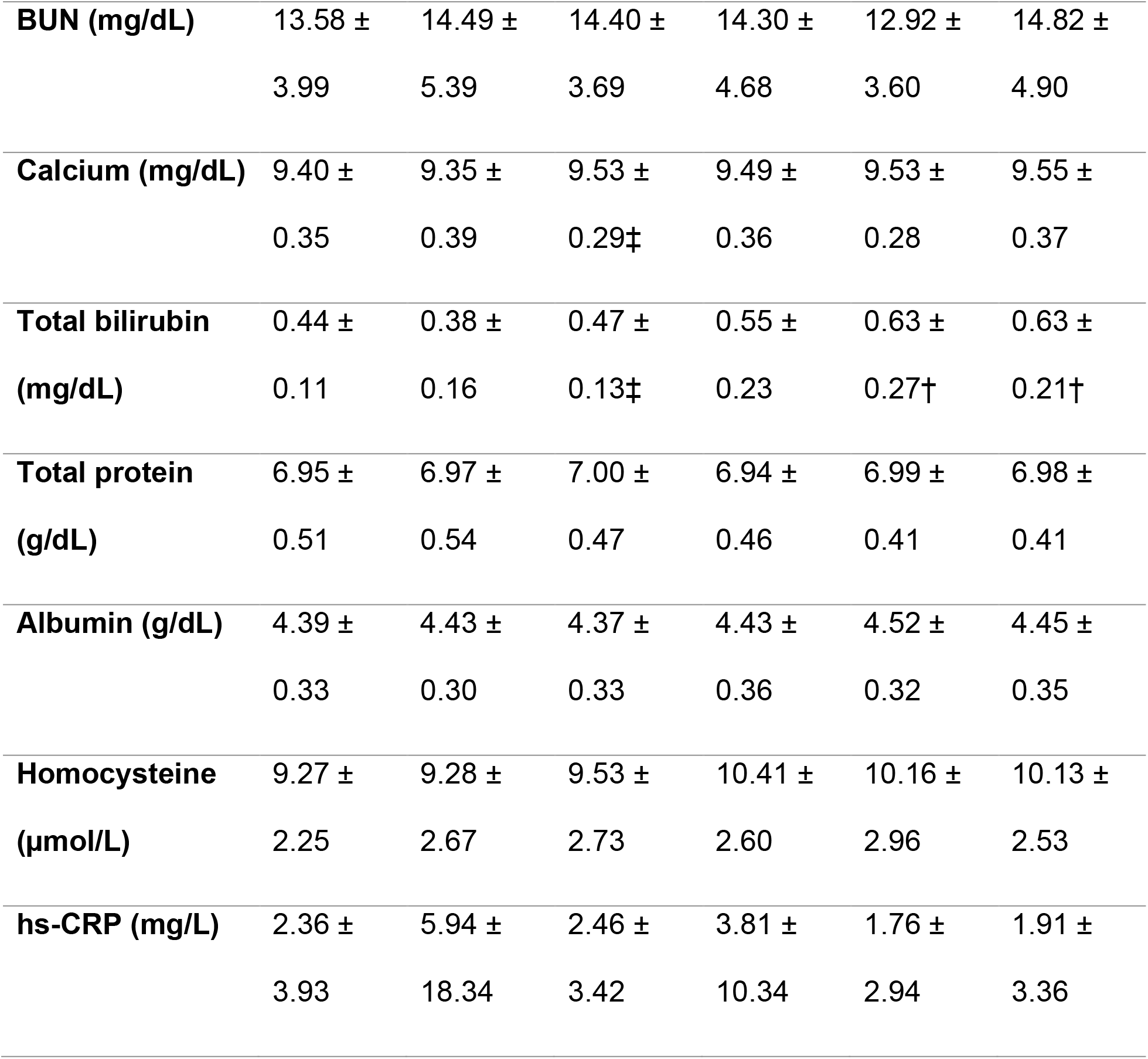
Clinical chemistry, hematology, and biomarker measures. Participants are grouped according to SC NR dose administered. BL represents Part 2 baseline assessment, followed by assessments at Day 30 and 90. Data are presented as mean ± SD. *p < 0.05 versus BL within the same dose group; ‡p < 0.05 versus Day 30 within the same dose group; †p < 0.05 versus the corresponding 50 mg group at the same timepoint

#### 3.2.3 Homocysteine

Blood homocysteine concentrations remained stable throughout the study, demonstrating no statistically significant changes (Table 4).

#### 3.2.4 hsCRP

Blood high-sensitivity c-reactive protein concentrations remained stable throughout the study, demonstrating no statistically significant changes (Table 4).

#### 3.2.5 Adverse Events

No serious AEs were reported. A total of 3 mild-to-moderate AEs occurred in the 100 mg group, which were: injection-site pain, hyperglycemia, and hypertension. The injection-site pain was attributed to likely contact with a superficial nerve during administration. The participant self-treated with a hot compress and Tylenol, and the event subsequently resolved. Regarding the hyperglycemia event, one participant in the 100 mg SC group had elevated glucose concentrations at baseline that persisted across subsequent assessments. The hypertension event was reported to the PI, who advised participant to follow up with their primary care physician. No AEs were reported among participants in the 50 mg group.

### 3.3 Whole-Blood NAD+

Whole-blood NAD+ concentrations remained relatively stable throughout the duration of the observation period, with no apparent increase in levels over time in either the 50 mg or 100 mg group. Concentrations were higher in the 100 mg group than in the 50 mg group across multiple timepoints, with significant between-group differences observed at baseline and Day 30 (both p <0.01), that remained significant at Day 90 (p < 0.05) (Figure 2A). This between-group difference was evident at the beginning of the at-home phase.

**Figure 2.**
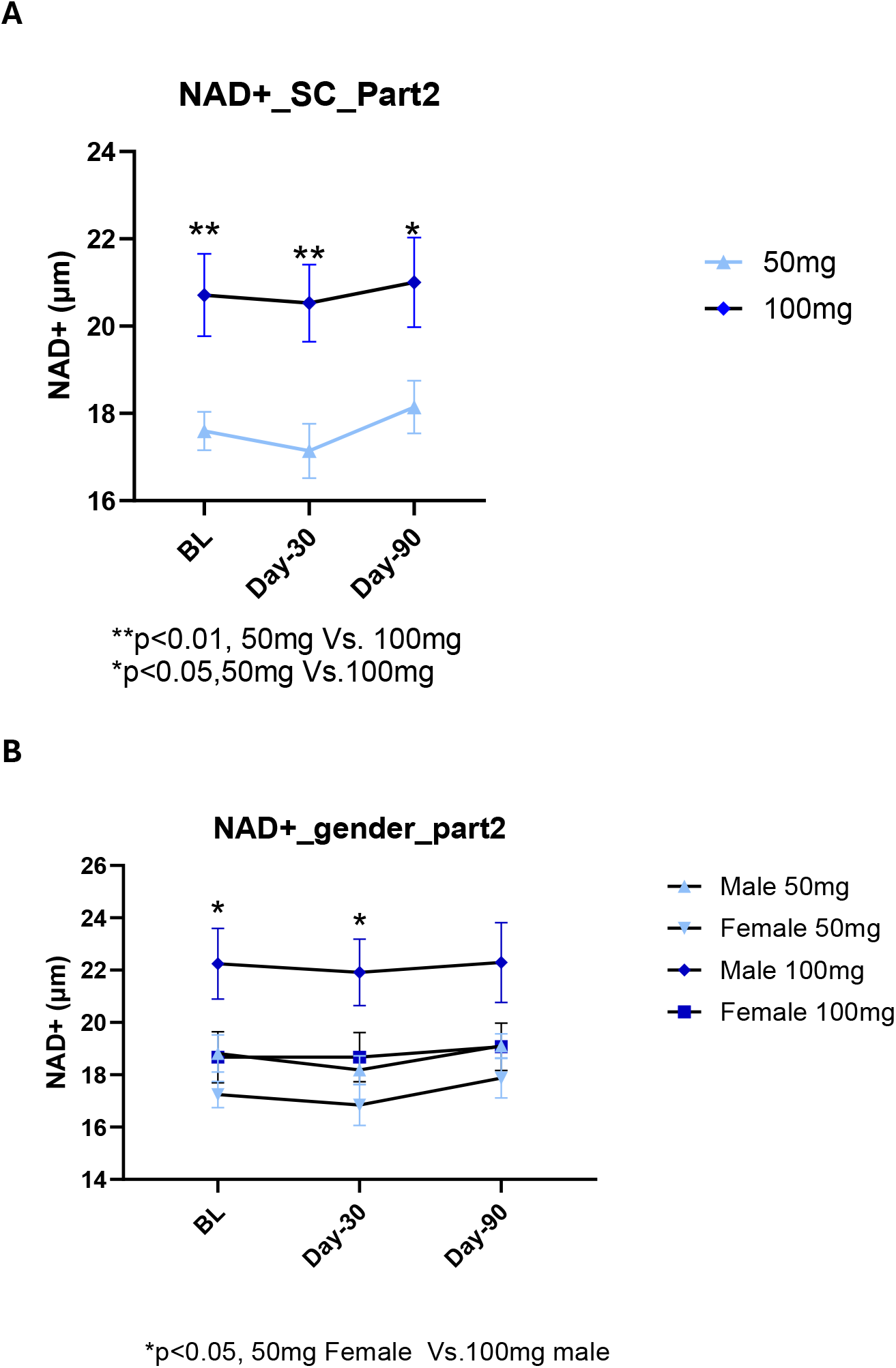
Whole blood NAD+ concentrations. Capillary whole blood NAD+ was measured by dried blood spots. NAD+ was measured at Part 2 baseline (BL), and after 30 and 90 days of at-home SC NR administration **A)** NAD+ concentrations stratified by dose group (50 mg and 100 mg) **B)** NAD+ concentrations further stratified by sex and dose. Data are presented as mean ± **SEM**. Asterisks denote statistically significant between-group comparisons: p < 0.05, and p<0.01.

When stratified by sex and dose, male participants in the 100 mg group exhibited the highest NAD+ concentrations. Levels were significantly higher in males receiving 100 mg compared with females receiving 50 mg at baseline and Day 30 (p<0.05); this difference was no longer significant at Day 90 (Figure 2B).

### 3.4 Mitochondrial Function

Mitochondrial bioenergetic outcomes were evaluated relative to study baseline (Visit 1). Analyses were conducted by 1) combining participants across 50 and 100 mg treatment groups to assess changes over time 2) stratifying by NR dose, and 3) stratifying by sex. For sex-stratified analyses, participants from both dose groups were pooled.

#### Pooled Analysis

When participants from the 50 and 100 mg groups were pooled, GlycoATP demonstrated a 15.05% reduction from baseline to Day 30 (p < 0.05), followed by a partial return toward baseline at Day 90 (Figure 3A). MitoATP increased directionally from baseline at Day 30, remaining elevated at Day 90, though these differences were not statistically significant (Figure 3B). Consistent with these changes, the MitoATP/GlycoATP ratio increased by 41.3% from baseline to Day 30 (p<0.01) and remained elevated at Day 90 (p<0.05) (Figure 3C)

**Figure 3.**
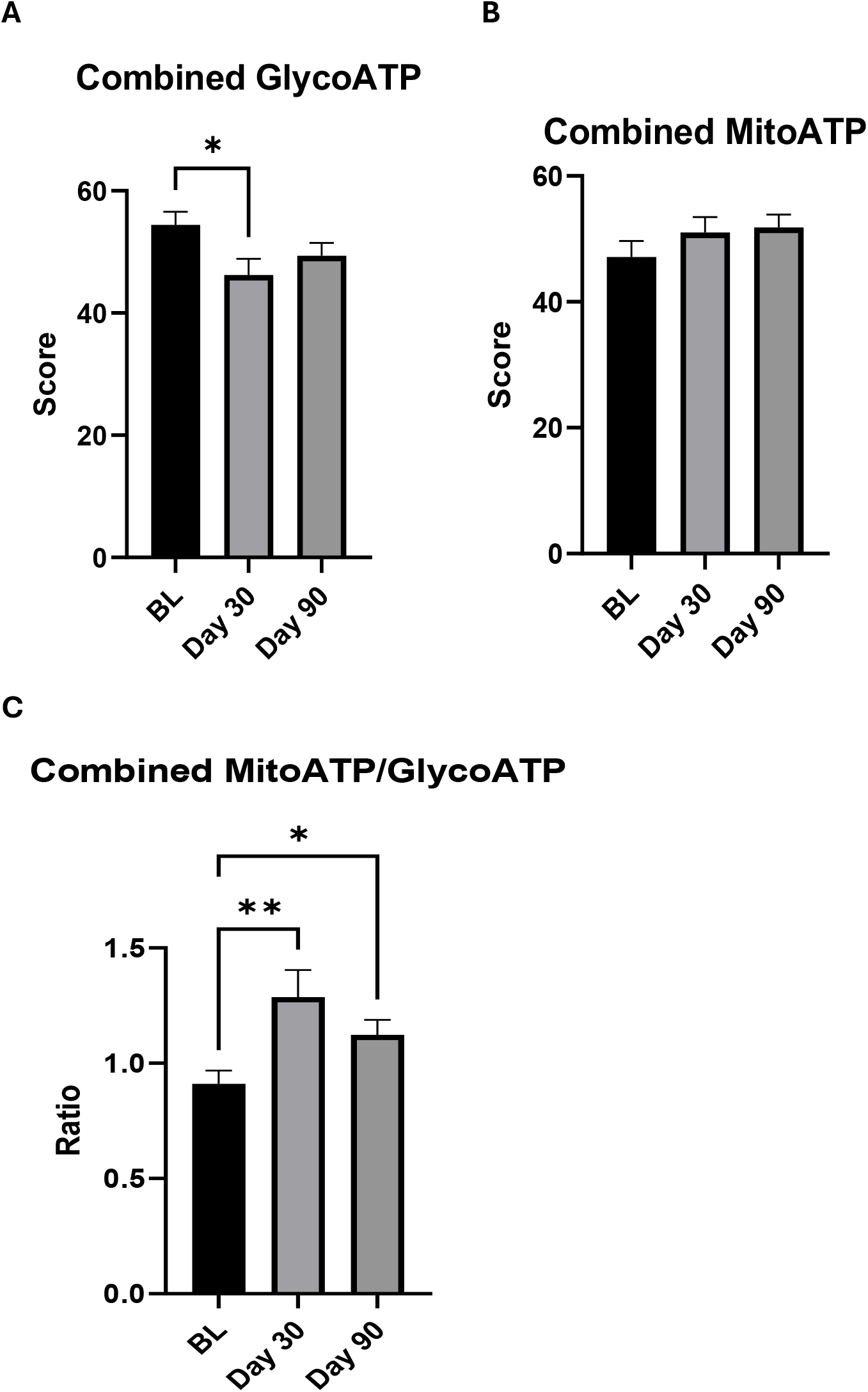
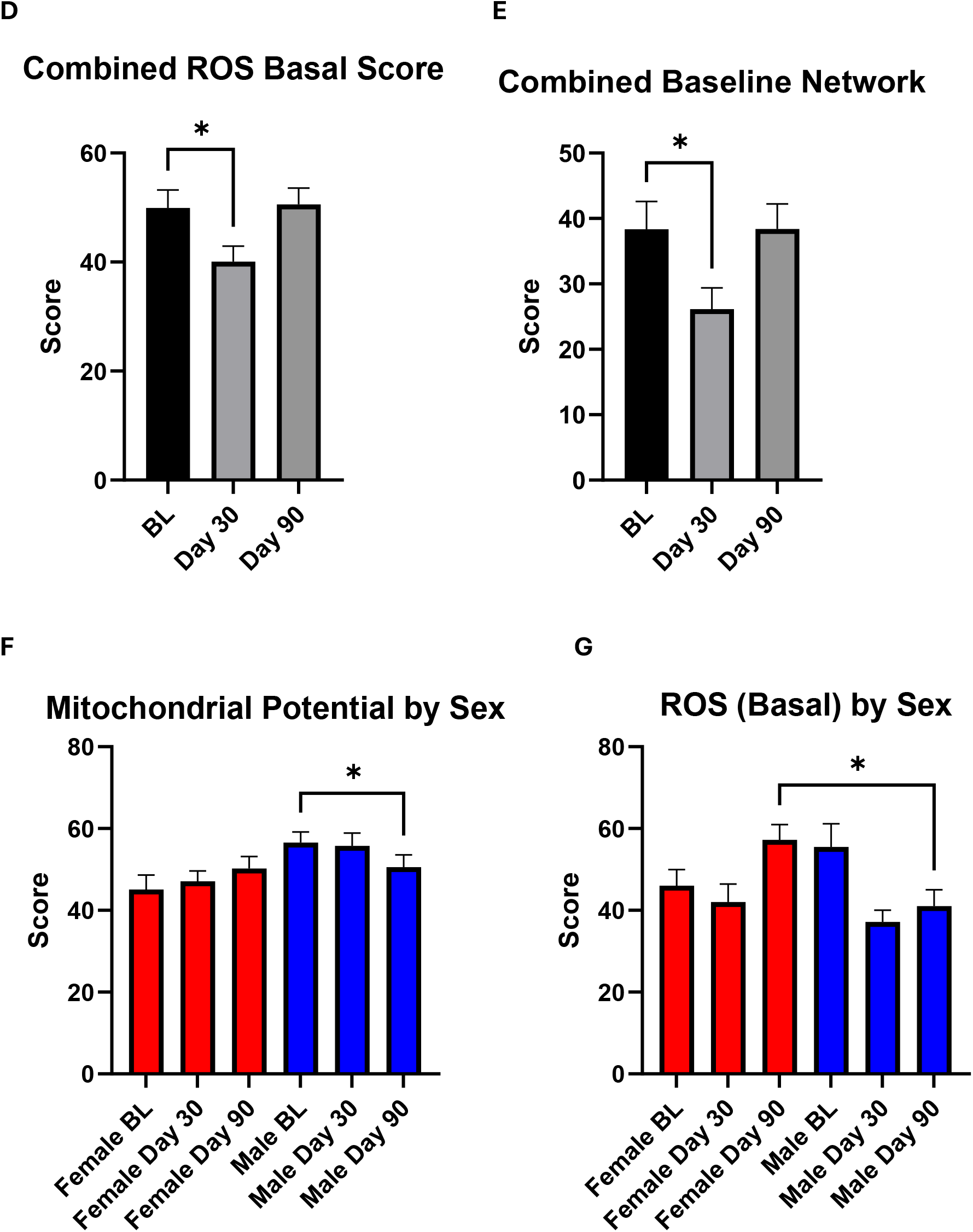
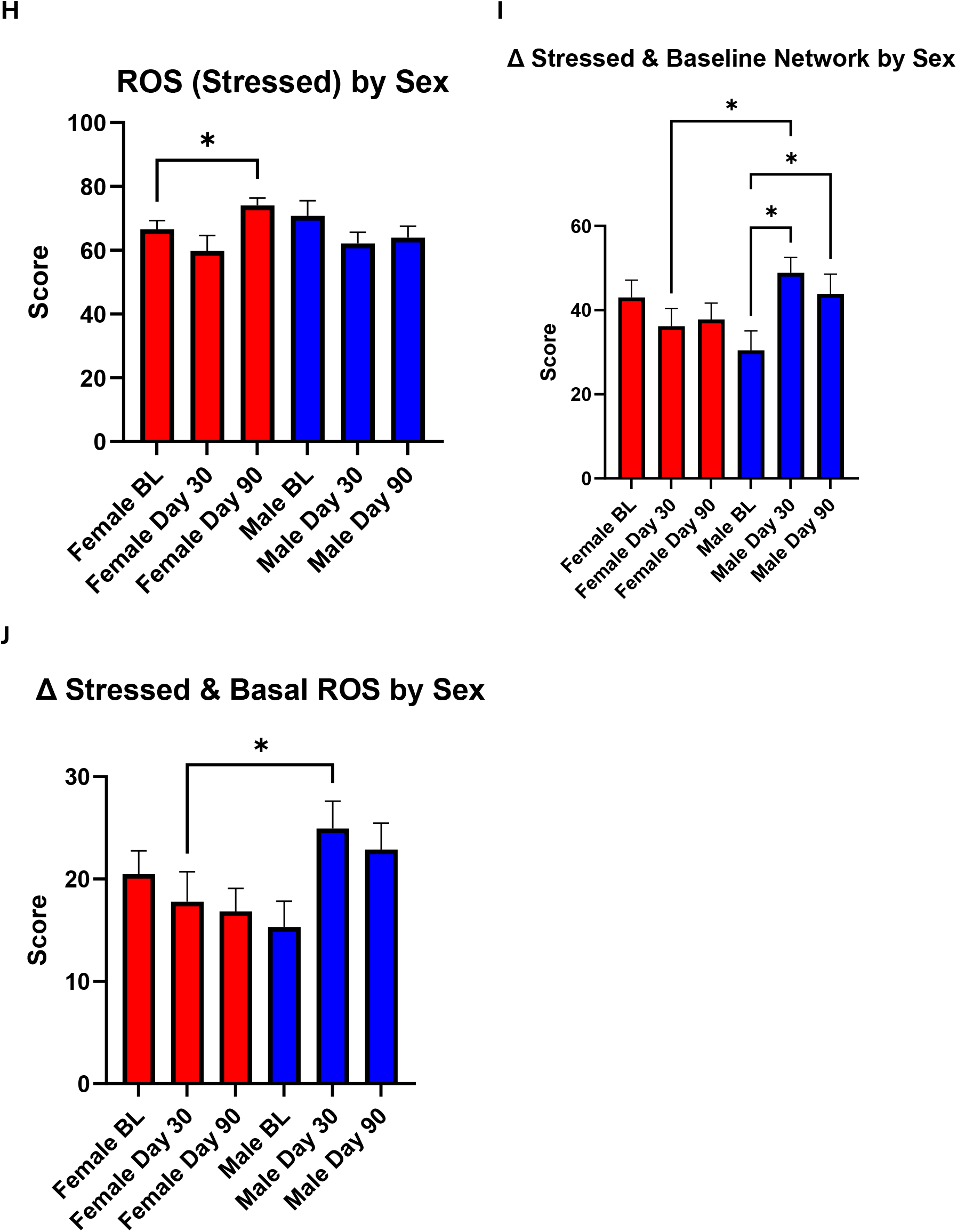
Mitochondrial parameters. Mitochondrial bioenergetic outcomes were assessed using the MeScreen platform at Part 1 baseline (BL), and after 30 and 90 days of at-home SC NR administration **A)** Glycolytic ATP production (GlycoATP) **B)** mitochondrial ATP production (MitoATP). **C)** MitoATP/GlycoATP ratio, **D)** basal reactive oxygen species (ROS), **E)** baseline mitochondrial network score **F)** mitochondrial potential by sex **G)** basal ROS stratified by sex, **H)** stressed ROS stratified by sex, **I)** change between stressed and baseline mitochondrial network scores stratified by sex, and **J)** change between stressed and basal ROS scores stratified by sex. For sex-stratified analyses, participants were pooled across NR dose groups. Data are presented as mean ± **SEM**. Asterisks denote statistically significant between-group comparisons: p < 0.05, and p<0.01.

Basal ROS significantly decreased from baseline to Day 30 (-19.8%, p <0.05), prior to returning to baseline at Day 90, while on the other hand, stressed ROS values were not statistically different at the three timepoints (Figures 3D and 3E). No significant changes were described for mitochondrial respiration, efficiency, or potential measures.

#### Dose-Stratified Analysis

When mitochondrial outcomes were analyzed separately according to NR dose, no significant changes from baseline to Day 30 or Day 90 in either the 50 mg or 100 mg groups were evident.

#### Sex-Stratified Analysis

Among male participants, mitochondrial potential was significantly decreased from baseline to Day 90, whereas mitochondrial potential appeared to increase directionally in a non-significant manner among females (Figure 3F). The stressed-basal mitochondrial network score also suggested sex-related differences; among males, this score was significantly higher at Day 30 and 90 compared to baseline (Figure 3I). At Day 30, female participants exhibited stressed-basal network scores that were significantly lower than their male counterparts.

With respect to ROS-related outcomes, females had significantly higher basal ROS at day 90 than males, and stressed ROS was higher at Day 90 compared to baseline among females (Figures 3G and 3H). In contrast, the stressed-basal ROS score was significantly higher among males compared with females at the day 30 timepoint (Figure 3J).

### 3.5 Urinary Oxidative Stress Biomarkers

Changes in urinary oxidative stress biomarkers relative to study baseline. Analyses were conducted by 1) combining participants across 50 and 100 mg treatment groups to assess changes over time 2) stratifying by NR dose, and 3) stratifying by sex. For sex-stratified analyses, participants from both dose groups were pooled.

#### Pooled analysis

When participants from both dose groups were analyzed in a pooled manner, 14 of the 16 biomarkers were significantly lower than baseline at one more timepoints.

Among markers of lipid oxidation, malondialdehyde, 8-iso-prostaglandin F2a, 11-β-prostaglandin F2α, and 15(R)-prostaglandin F2α were significantly lower at Day 30 compared with baseline (all p< 0.05; Figures 4A-D). However, these differences did not remain statistically significant at Day 90, with concentrations generally returning toward baseline values.

**Figure 4.**
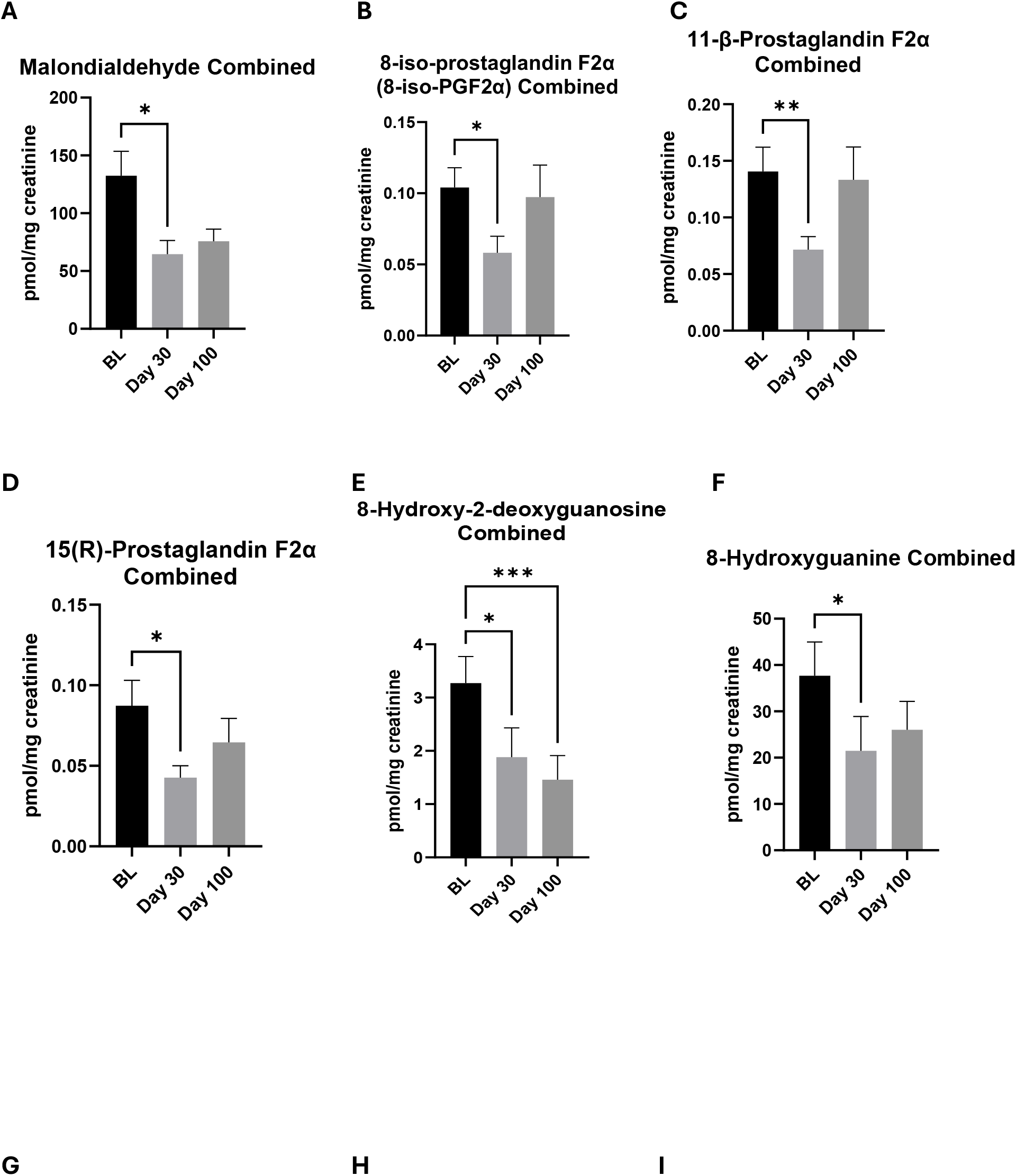
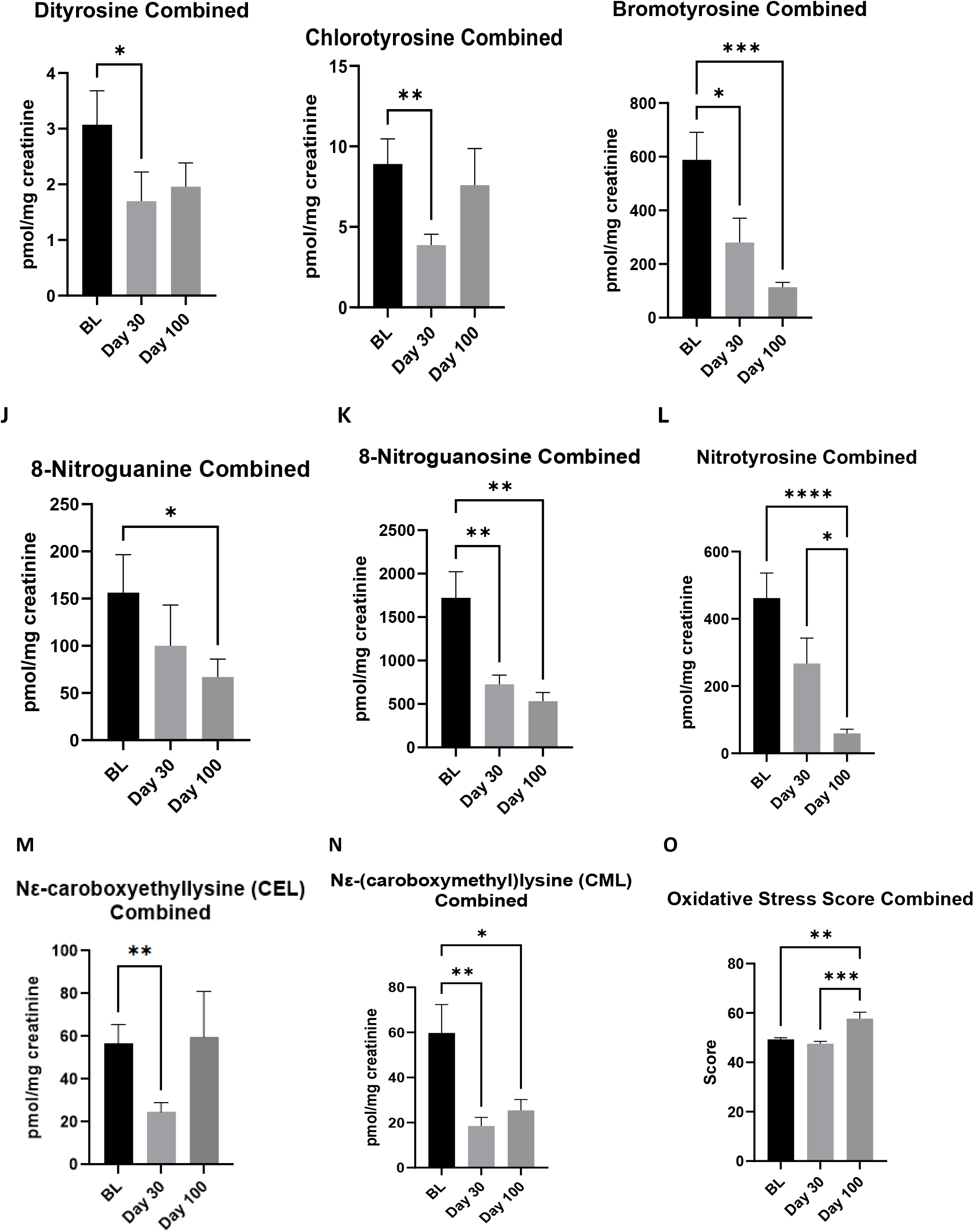
Urinary oxidative stress biomarkers in the pooled analysis. Urinary oxidative stress biomarkers were assessed at Part 1 baseline (BL), and after 30 and 90 days of at-home SC NR administration. Participants from both 50 and 100 mg groups were pooled for analysis. Biomarkers that demonstrated at least one statistically significant change are included: **A)** malondialdehyde, **B)** 8-iso-prostaglandin F2α, **C)** 11-β-prostaglandin F2α, **D)** 15(R)-prostaglandin F2α, **E)** 8-hydroxy-2′-deoxyguanosine, **F)** 8-hydroxyguanine, **G)** dityrosine, **H)** chlorotyrosine, **I)** bromotyrosine, **J)** 8-nitroguanine, **K)** 8-nitroguanosine, **L)** nitrotyrosine, **M)** Nε-carboxyethyllysine, **N)** Nε-(carboxymethyl)lysine, and **O)** composite oxidative stress score. Data are presented as mean ± **SEM**. Asterisks denote statistically significant comparisons: p < 0.05, and p<0.01 and p <0.001.

Among markers of DNA/RNA oxidation, 8-hydroxy-2-deoxyguanosine concentrations were significantly lower at both Day 30 and Day 90 in comparison to baseline (p <0.05 Day 30 vs baseline, p <0.01 Day 90 vs baseline; Figure 4E). 8-Hydroxyguanine fell significantly at Day 30 (p<0.05), but returned toward baseline at Day 100 (Figure 4F). In contrast, 8-Hydroxyguanosine levels were not significantly altered.

Oxidative markers in relation to protein oxidation, nitrative stress, and advanced glycation end-products were also modified throughout the study period. Dityrosine, chlorotyrosine, Nε -carboxyethyllysine concentrations were lower at Day 30 (p < 0.05, 0.01, 0.01), but increased toward baseline by Day 90 (Figures 4G, H, M). On the other hand, bromotyrosine, 8-nitroguanosine, Nε-(carboxymethyl)lysine, demonstrated more sustained responses, with concentrations statistically lower at Days 30 and 90 relative to baseline (p < 0.01, 0.01, 0.05; Figures 4I, K N). Similarly, nitrotyrosine exhibited a progressive decline over time, reaching statistical significance at Day 90 relative to both baseline (p<0.001) and Day 30 (p<0.05) (Figure 4L). 8-nitroguanine was numerically lower at Day 30, and only reached statistical significance at Day 90 relative to baseline (Figure 4J).

The composite oxidative stress score changed by 17% from baseline to Day 90 (p <0.05; Figure 4O).

#### Dose-stratified analysis

Dose-stratified analyses yielded generally comparable directional changes in the 50 mg and 100 mg groups, although substantially fewer comparisons reached statistical significance.

In the 50 mg group, 11-β-Prostaglandin F2α and 8-hydroxy-2-deoxyguanosine decreased significantly from baseline to Day 30 (p<0.05; Figure 5A, B). Nitrotyrosine levels fell over time, with significant reductions observed at Day 30 and Day 90 (both p <0.05) in comparison to baseline (Figure 5C).

**Figure 5.**
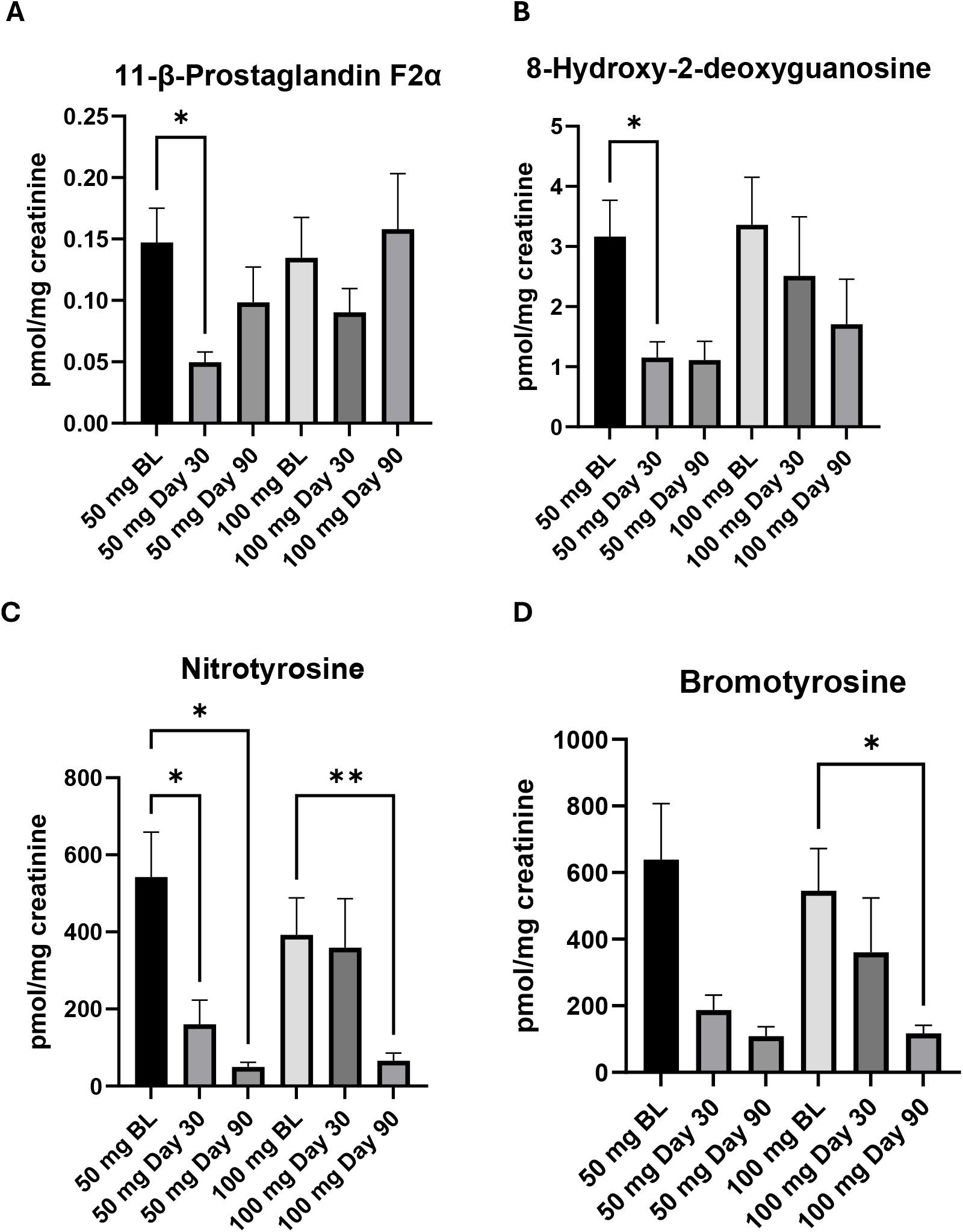
Dose-stratified changes in urinary oxidative stress biomarkers. Longitudinal changes in oxidative stress biomarkers were assessed separately in participants receiving 50 or 100 mg subcutaneous NR. Panels depict biomarkers demonstrating at least one statistically significant within-dose longitudinal change: **A)** 11-β-prostaglandin F2α, **B)** 8-hydroxy-2′-deoxyguanosine, **C)** nitrotyrosine, and **D)** bromotyrosine. Data are presented as mean ± **SEM**. Asterisks denote statistically significant comparisons: p < 0.05, and p<0.01 and p <0.001.

In the 100 mg group, bromotyrosine and nitrotyrosine levels were significantly reduced at Day 90 compared to baseline (p <0.05, 0.01; Figure 5C, D).

For both dose groups, additional oxidative stress biomarkers demonstrated numerical reductions at certain follow-up assessments, but these changes failed to reach statistical significance.

#### Sex-stratified analysis

When participants from both groups were combined and subsequently stratified by sex, significant longitudinal changes were observed in both female and male participants, although the pattern of statistically significant findings varied between sexes.

In female participants, bromotyrosine concentrations declined progressively over the study period and were significantly lower at Day 90 compared with baseline (p < 0.01; Figure 6A). Chlorotyrosine concentrations were significantly lower at Day 30 compared with baseline (p < 0.05), but subsequently increased toward baseline by Day 90 (Figure 6B). Markers of nitrative stress were also altered; 8-nitroguanosine and nitrotyrosine concentrations decreased over time, and were significantly lower at Day 90 compared with baseline (p<0.05; Figure 6C, D). Among advanced glycation markers, Nε-(carboxymethyl)lysine was significantly reduced at Day 30 compared with baseline (p<0.05), but subsequently increased toward baseline by Day 90 (Figure 6E). The composite oxidative stress score decreased at Day 30, but increased significantly from Day 30 to Day 90 (p<0.05; Figure 6F). The remaining oxidative stress markers remained statistically unaltered, though several demonstrated numerical reductions at one or more timepoints.

**Figure 6.**
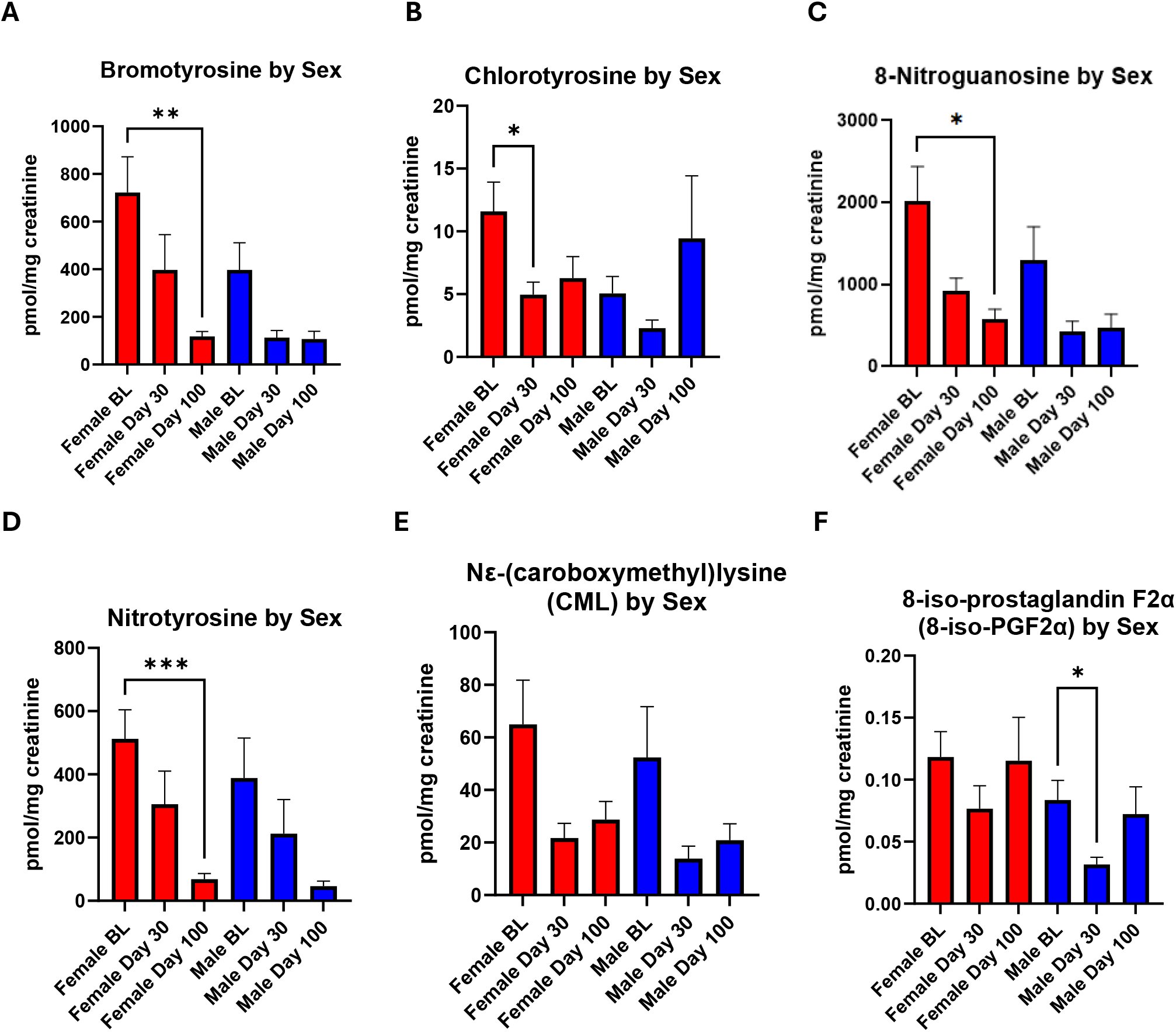
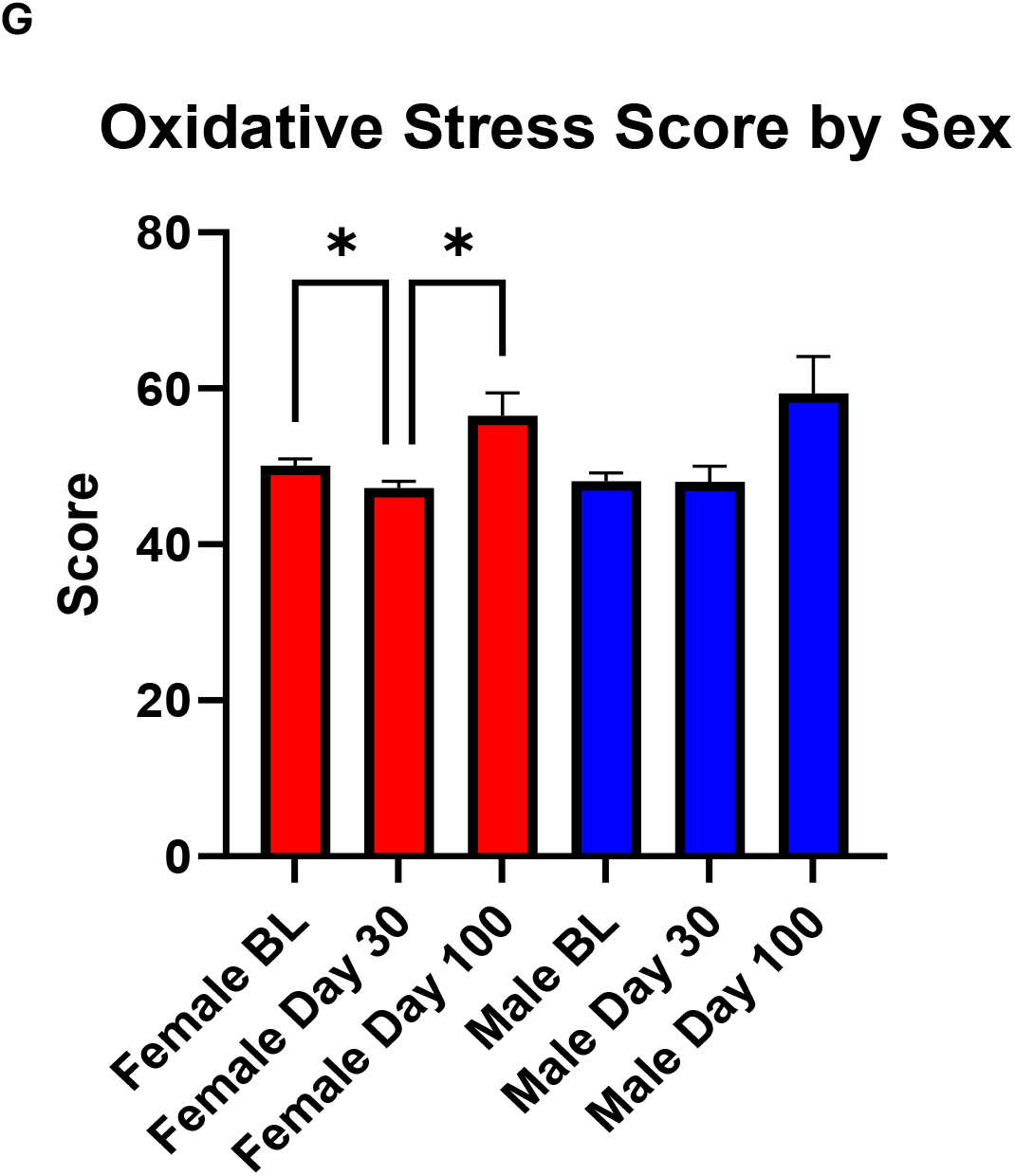
Sex-stratified changes in urinary oxidative stress biomarkers. Participants were pooled across NR dose groups and stratified by sex. Panels depict biomarkers demonstrating at least one statistically significant longitudinal change within sex: **A)** bromotyrosine, **B)** chlorotyrosine, **C)** 8-nitroguanosine, **D)** nitrotyrosine, E) Nε-(carboxymethyl)lysine, **F)** 8-iso-prostaglandin F2α, and **G)**. composite oxidative stress score Data are presented as mean ± SEM. Asterisks denote statistically significant comparisons: p < 0.05, and p<0.01 and p <0.001.

In male participants, 8-iso-prostaglandin F2α was significantly lower at Day 30 compared with baseline (p<0.05), but subsequently increased by Day 90 (Figure 6G). No other oxidative stress markers demonstrated a significant change across the study period.

### 3.7 Participant-Reported Outcomes

Participant-Reported Outcomes (PROs) were analyzed by 1) combining participants across 50 and 100 mg treatment groups to assess changes over time 2) stratifying by NR dose, and 3) stratifying by sex. For sex-stratified analyses, participants from both dose groups were pooled.

#### 3.7.1 Fatigue

##### Pooled analysis

PROMIS Fatigue scores decreased over the study period, with scores significantly lower at Day 90 compared to baseline (p < 0.01; Figure 7A).

**Figure 7.**
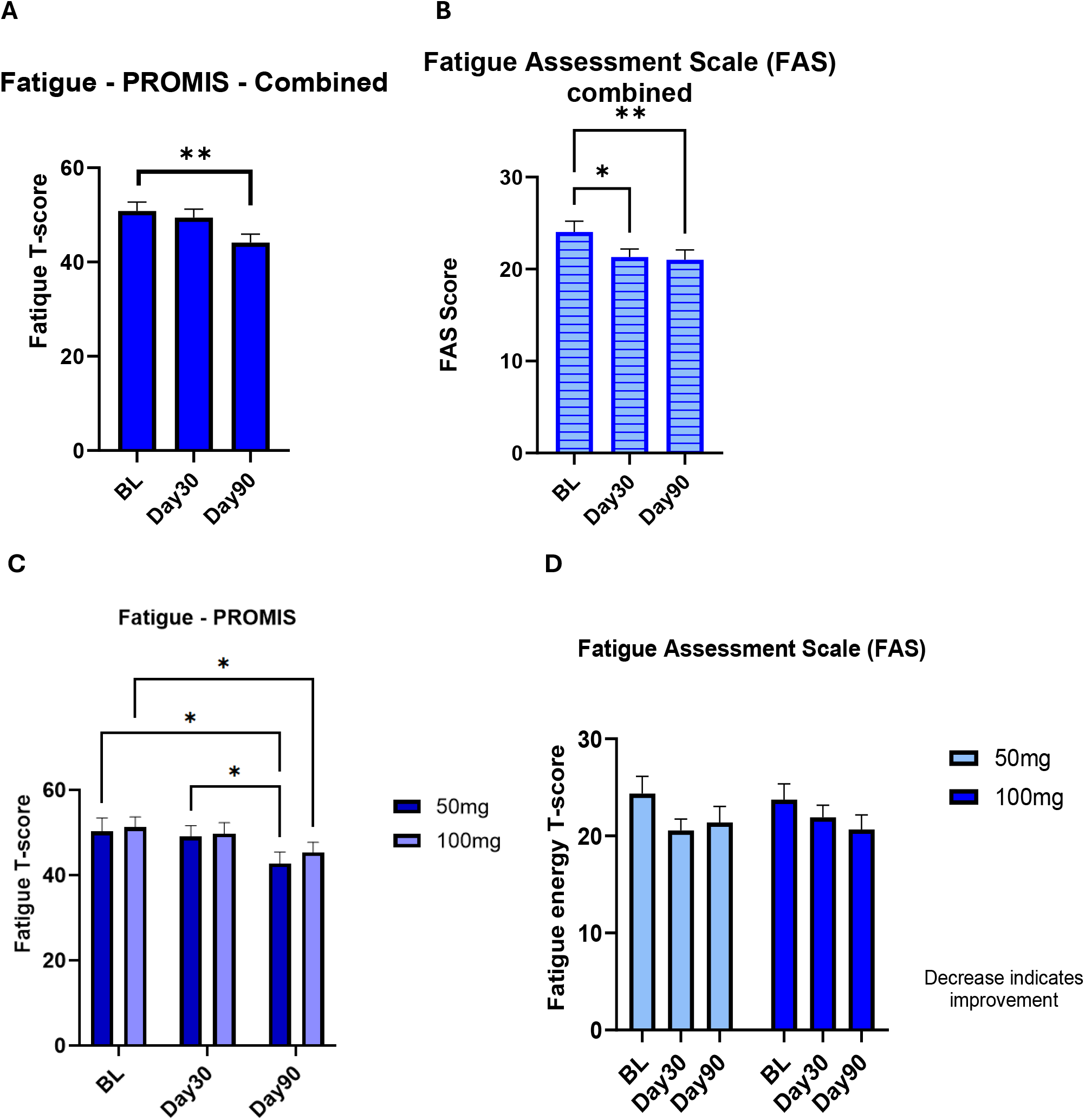
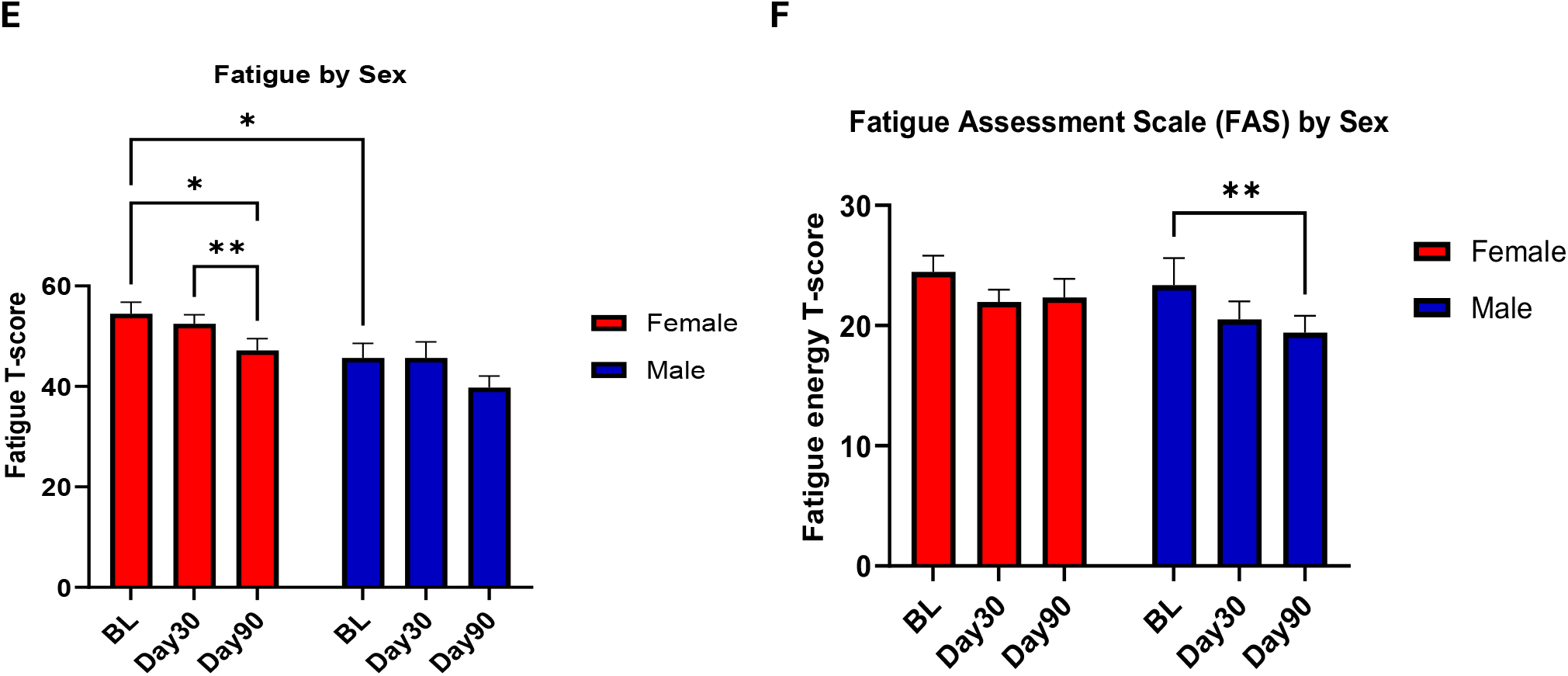
Participant-reported fatigue during at-home NR administration. Mean scores are shown at baseline, Day 30, and Day 90. Pooled analysis: **A)** PROMIS Fatigue T-score and **B)** Fatigue Assessment Scale (FAS). Dose-stratified analysis: **C)** PROMIS Fatigue T-score and **D)** FAS, stratified by 50 mg and 100 mg NR dose. Sex-stratified analysis: **E)** PROMIS Fatigue T-score and **F)** FAS, stratified by sex. Higher scores indicate greater fatigue. Data are presented as mean ± SEM. Asterisks denote statistically significant comparisons: *p* < 0.05, **p** < 0.01, and ***p*** < 0.001.

FAS scores also showed reductions, with Day 30 and Day 90 scores both lower than baseline levels (p <0.05, 0.01; Figure 7B).

##### Dose-stratified analysis

When stratified by dose, both the 50 mg and 100 mg groups demonstrated significant ly lower PROMIS Fatigue T-scores at Day 90 compared with baseline (p <0.05; Figure 7C).

In contrast, no statistically significant changes were observed in FAS scores, when stratified by dose (Figure 7D).

##### Sex-stratified analysis

When stratified according to sex, female participants had higher levels of fatigue at baseline, as measured by the PROMIS Fatigue instrument, compared to their male counterparts (p<0.05). Day 90 scores were significantly lower than Day 30 and baseline scores among females (p<0.01, 0.05), whereas no significant changes were observed across timepoints for males (Figure 7E)

In contrast, FAS scores were significantly lower at Day 90 vs. baseline (p<0.01) in males; no significant within-group changes were noted in female participants (Figure 7F).

#### 3.7.2 Sleep Disturbance and Sleep-Related Impairment

##### Pooled analysis

No significant changes in sleep disturbance were observed across any of the assessed timepoints (Figure 8A).

**Figure 8.**
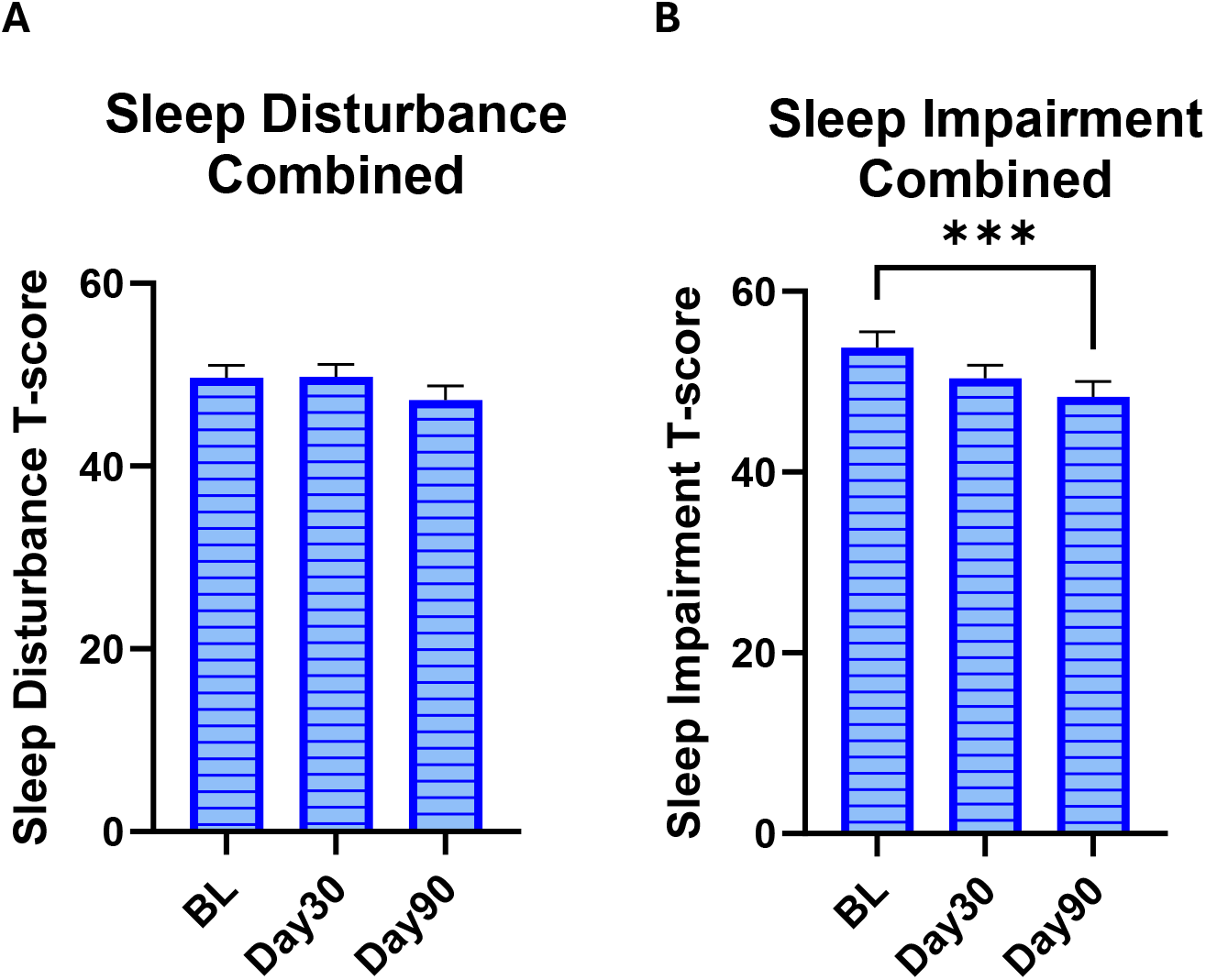
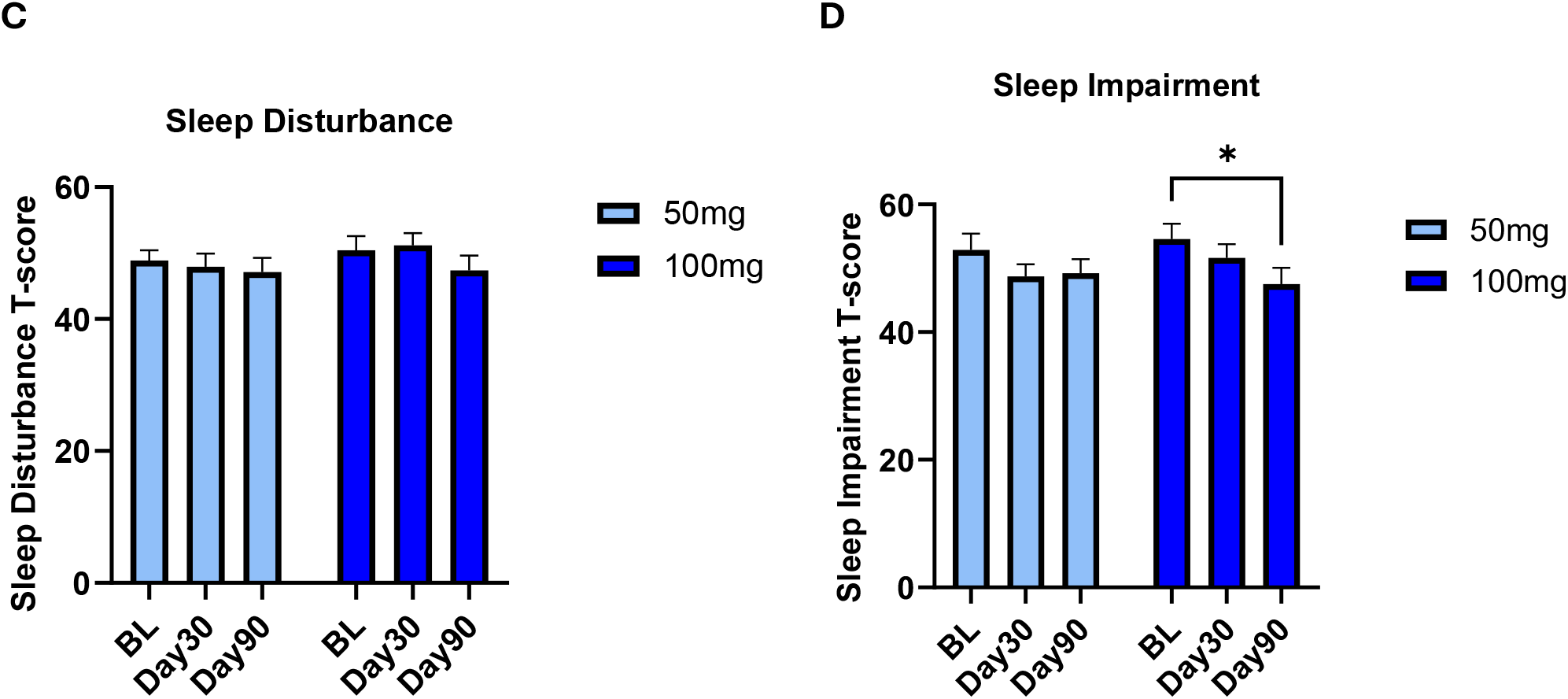
Participant-reported sleep outcomes during at-home NR administration. Mean scores are shown at baseline, Day 30, and Day 90. Pooled analysis: **A)** PROMIS Sleep Disturbance T-score and **B)** PROMIS Sleep-Related Impairment T-score. Dose-stratified analysis: **C)** Sleep Disturbance T-score and **D)** Sleep-Related Impairment T-score, stratified by 50 mg and 100 mg NR dose. Higher scores indicate greater sleep disturbance or sleep-related impairment. Data are presented as mean ± SEM. Asterisks denote statistically significant comparisons: *p* < 0.05, **p** < 0.01, and ***p*** < 0.001.

Sleep-related impairment scores were significantly lower at Day 90 vs. baseline (p<0.001; Figure 8B).

##### Dose-stratified analysis

Sleep disturbance T-scores declined directionally in the 50 mg group, although no comparisons achieved statistical significance. In the 100 mg group, sleep disturbance tended to be lower at Day 90 compared to Day 30 (p = 0.07; Figure 8C).

Concerning the sleep-related impairment assessment, the 100 mg group demonstrated statistically significant reductions in T-scores at Day 90 relative to baseline (p <0.05). In the 50 mg group, the scores showed progressive directional reductions, though none of the differences achieved statistical significance (Figure 8D).

##### Sex-stratified analysis

No significant changes in sleep disturbance were observed across any of the assessed timepoints, when stratified by sex.

No significant changes in sleep disturbance were observed across any of the assessed timepoints, when stratified by sex.

#### 3.7.3 Quality of Life – Global Mental and Physical Health

##### Combined analysis

GMH T-scores increased significantly by Day 90 compared to baseline (p <0.05; Figure 9A).

**Figure 9.**
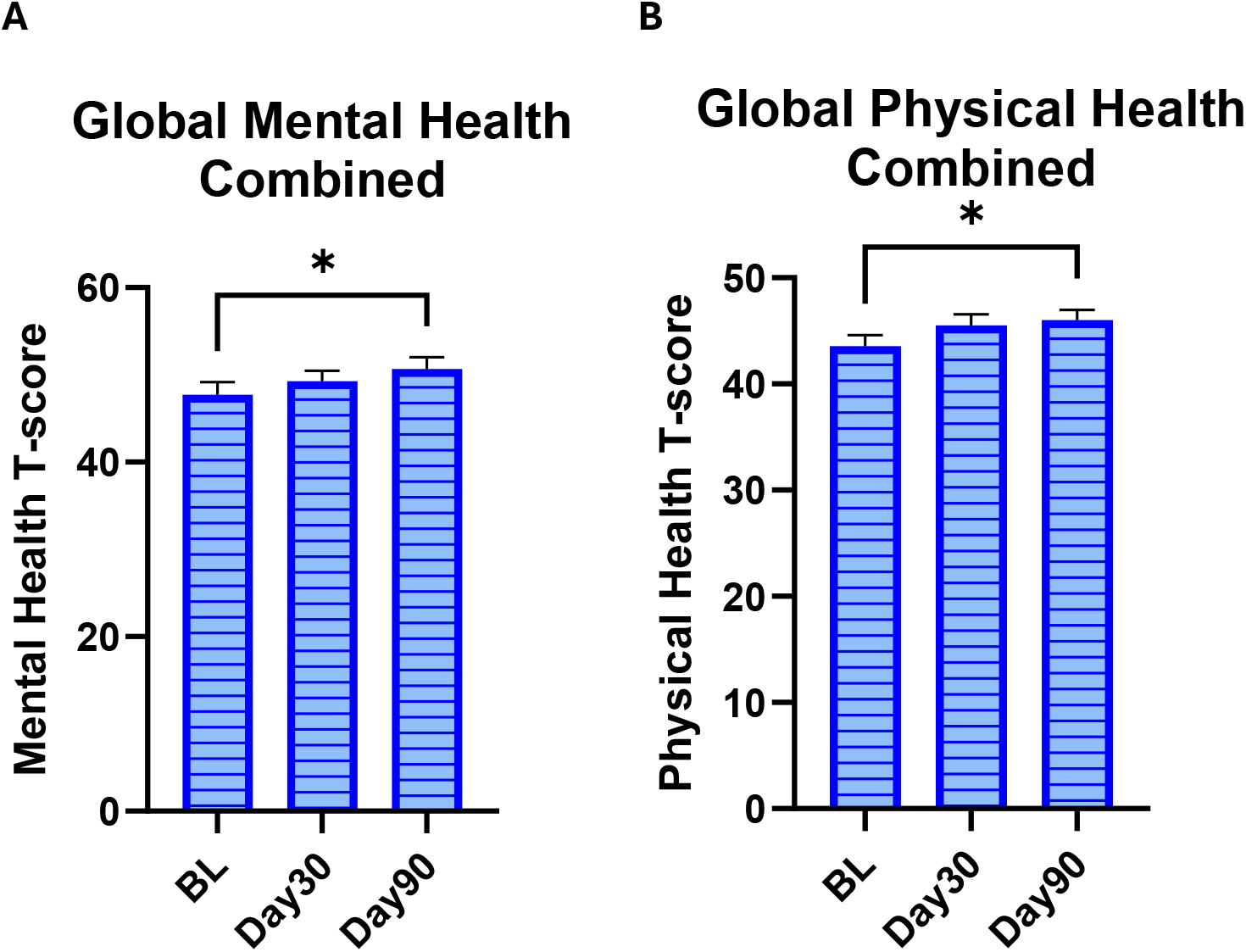
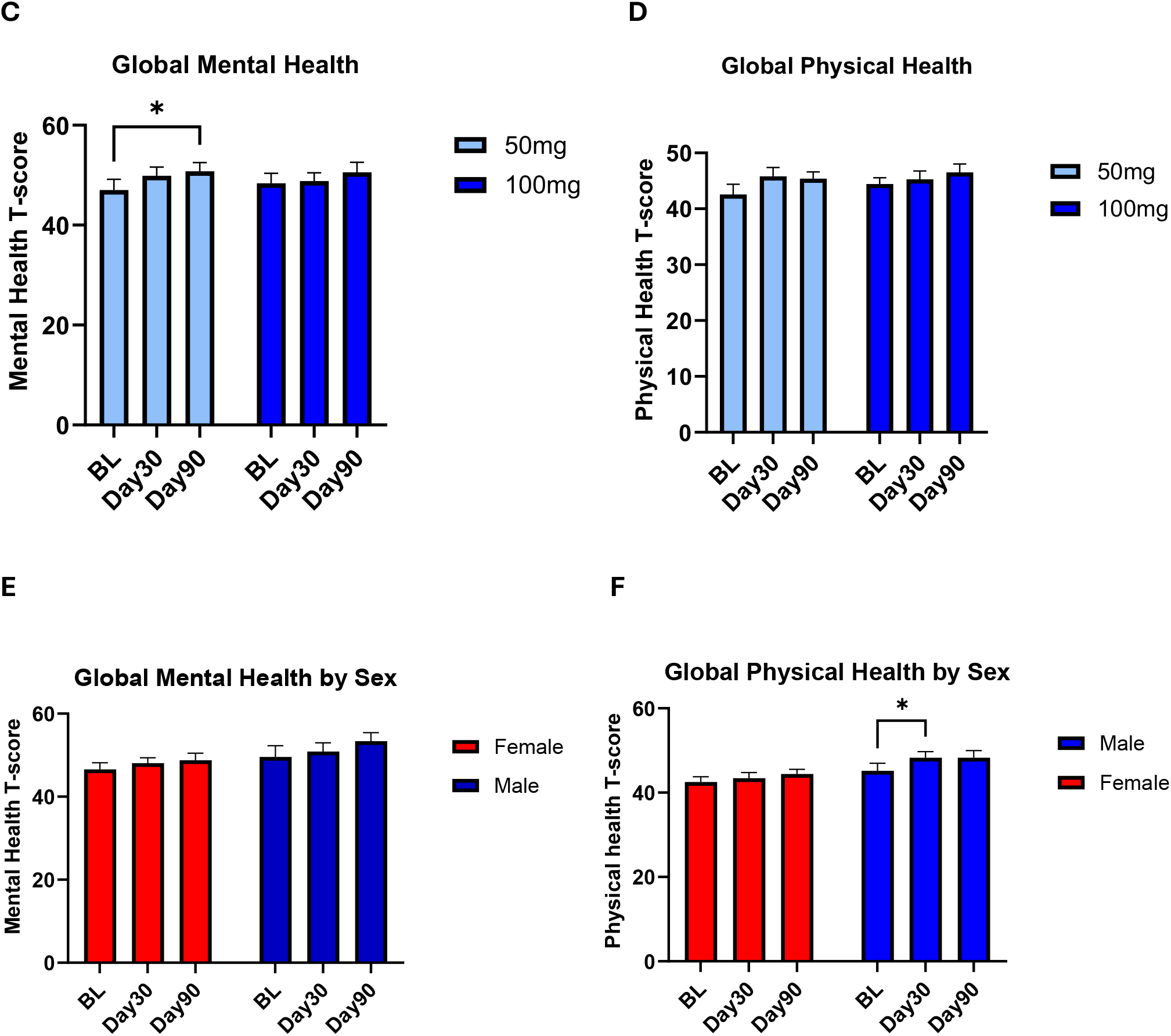
Participant-reported global health during at-home NR administration. Mean scores are shown at baseline, Day 30, and Day 90. Pooled analysis: **A)** PROMIS Global Mental Health T-score and **B)** PROMIS Global Physical Health T-score. Dose-stratified analysis: **C)** Global Mental Health T-score and **D)** Global Physical Health T-score, stratified by 50 mg and 100 mg NR dose. Sex-stratified analysis: **E)** Global Mental Health T-score and **F)** Global Physical Health T-score, stratified by sex. Higher scores indicate better perceived health. Data are presented as mean ± SEM. Asterisks denote statistically significant comparisons: *p* < 0.05, **p** < 0.01, and ***p*** < 0.001.

GPH T-Scores also increased significantly by Day 90 compared to baseline (p<0.05; Figure 9B).

##### Dose-stratified analysis

GMH T-scores showed progressive directional increases in both 50 mg and 100 mg groups. Day 90 scores were significantly higher compared to baseline in the 50 mg group, whereas no significant differences were observed in the 100 mg group (Figure 9C).

GPH T-scores were not significantly different at any timepoint for either dose group, although scores at Day 30 tended to be higher than baseline in the 50 mg group (p = 0.07; Figure 9D).

##### Sex-stratified analysis

No significant changes in GMH were observed across any of the assessed timepoints, when stratified by sex (Figure 9E).

GPH scores were significantly increased at Day 30 compared to baseline among male participants (p<0.05); no significant changes were observed for female participants (Figure 9F).

### 3.8 Self-Reported Skin Outcomes

Participant-reported skin outcomes were assessed across four domains at baseline, Day 30, and Day 90 using open-ended questionnaire responses, which were categorized as improvement, no change, worsening, other/neutral change, or unclear.

The clearest longitudinal pattern observed was a progressive increase in the frequency of self-reported skin improvements for “facial skin”, “facial changes noticed by others”, and “body skin” domains (Table 5). Reports of other favorable physical appearance changes were less consistent over time.

**Table 5.** Self-reported skin improvement by domain.

| Domain | Day 1 | Day 30 | Day 90 |
| --- | --- | --- | --- |
| Facial skin | 11.4% (4/35) | 27.8% (10/36) | 36.8% (14/38) |
| Facial changes noticed by others | 9.1% (3/33) | 17.6% (6/34) | 23.7% (9/38) |
| Body skin | 8.8% (3/34) | 9.1% (3/33) | 16.7% (6/36) |

Of note, a small proportion of participants (3-8.3%) reported unfavorable skin changes, including dryness, itching, and breakouts. In sum, the descriptive findings suggest an increasing frequency of perceived skin improvements over the study period, while changes in other aspects of physical appearance were less consistent.

## 4. Discussion

Interest in strategies to modify NAD+ parenterally has outpaced clinical evidence supporting their use, particularly for routes suited for self-administration. While oral administration of NR and other NAD+ precursors have generally been shown to be safe and well-tolerated, recent data have suggested route-related differences (e.g., SC, IM, IV) in the safety of NR^31^, precluding direct extrapolation of oral pharmacokinetics and safety assumptions to parenteral routes of administration. To our knowledge, the present study represents the first to evaluate repeated self-administration of SC NR over an extended period. The principal finding was that at-home SC injections of 50 or 100 mg of NR three times weekly over 90 days were safe and well-tolerated, with no dose-related safety signal across vital signs, clinical health and safety parameters, or adverse events. Interestingly, whole-blood NAD+ concentrations remained stable during the 90-day at-home administration period, despite continued exposure to NR. In parallel, exploratory analyses identified longitudinal changes in selected measures of mitochondrial bioenergetics, urinary oxidative stress, and participant-reported outcomes – all of which were altered directionally in a favorable manner. Of note, these benefits occurred without a coinciding increase in whole-blood NAD+, with little evidence of a dose-dependent response.

The safety findings reported herein extend upon the short-term observations from Part 1 to an unsupervised, at-home setting. Vital signs remained stable, and no meaningful changes occurred in hematological and chemical parameters; alterations were quantitatively minor and not clinically relevant. Three AEs were reported, consisting of injection-site pain, isolated increases in blood pressure, and plasma glucose. The participant who presented with marked hyperglycemia likely had undiagnosed type-2 diabetes at baseline which was not adequately screened prior to enrollment; a causal relationship to NR cannot be established. Nevertheless, effects of NAD+ precursors on glucose homeostasis have been reported in select previous studies. For instance, Lapatto et al. observed a modest, but significant increase in fasting glucose with oral 1000 mg NR/day ^37^. In addition, niacin (nicotinic acid) is known to induce fasting hyperglycemia at high doses in some individuals, and as a result, is contraindicated in those with impaired fasting glucose^38^. The preclinical literature on oral NR is also mixed; NR at doses of 400-500 mg/kg/day improved glucose tolerance and mediated an overall beneficial metabolic effect in animals treated with a high-fat diet^12^, however, glucose homeostasis was disturbed when a dose of 1000 mg/kg/day was used^39^. Thus, there may be individual and dose-related responses to NAD+ precursor utilization that could have either positive or negative metabolic consequences, thereby necessitating the need for future NR studies to screen participants for impaired glucose homeostasis prior to enrollment. Taken together, the absence of clinically meaningful changes across multiple health indices supports the overall safety and tolerability of 50 mg and 100 mg of SC NR, and is consistent with the favorable tolerability profile reported for IV NR in two previous clinical studies^29,30^. Given the modest sample size, these findings do not exclude the possibility of uncommon, idiosyncratic AEs or subtle safety differences between doses or administration routes.

In Part 1, three consecutive daily injections increased whole-blood NAD+ by Day 3. However, herein, no further increase was observed over 90 days of at-home administration at either dose. Several factors may explain these findings. First, daily oral NR supplementation requires about 10-14 days to achieve a new steady-state whole-blood NAD+ concentration^25^. The 3x/week schedule may therefore have been insufficient to sustain elevated NAD+ between doses, and more frequent administrations may be required to induce progressive increases in whole blood NAD+. Second, whole blood NAD+ may not provide a complete representation of NAD+ metabolism across tissues. Whole blood measurements are predominantly reflective of erythrocyte NAD+, whereas the NAD+ responsiveness to NR may differ substantially across cell and tissue types. Therefore, the absence of a measurable, sustained increase in whole blood NAD+ does not preclude the presence of NAD+-related metabolic effects in other tissues and organs.

The relatively modest increase in whole blood NAD+ during Part 1, followed by no further change in Part 2 may also reflect differences in NR metabolism between parenteral and oral routes. When orally administered, a significant portion of NR is converted to other NAD+ precursors and related metabolites during first-pass metabolism. Briefly, this process involves the activity of BST1 (bone marrow stromal cell antigen 1; also recognized as ADP-ribosyl cyclase 2 and CD157), which hydrolyzes NR to form nicotinamide^26,28^. A meaningful fraction of the resultant nicotinamide is then further biotransformed to nicotinic acid, catalyzed by nicotinamidase enzymes secreted by the gut microbiota^40^. Therefore, nicotinic acid and nicotinamide derived from NR may also subsequently contribute to NAD+ biosynthesis through salvage and Preiss-Handler pathways, such that the systemic effects of oral NR likely reflect exposure to a mixture of NR and other related B3 vitamin compounds, rather than intact NR alone.

Parenteral administration bypasses gastrointestinal and microbial digestion and therefore may potentially provide a more direct and predictable route of systemic NR delivery. In principle, SC or IM administration may increase the intact NR exposure, by reducing variability related to gastrointestinal metabolism, and permit more rapid systemic availability, in comparison to oral administration. Nevertheless, these theoretical advantages remain putative, as this study did not directly compare oral and SC NR pharmacokinetics, or assess tissue-specific NAD+ concentrations. Preclinical studies evaluating intravenous administration have suggested that it facilitates greater uptake of intact NR in comparison to orally administered NR^41^.

The urinary oxidative stress panel provided exploratory evidence of longitudinal biological changes. Statistically significant reductions were detected for 14 of the 16 measured oxidative stress markers at at least one timepoint relative to baseline, and all showed directional improvement, when all participants were pooled. Several lipid peroxidation markers were reduced at Day 30, but returned toward baseline by Day 90. In contrast, selected markers of DNA/RNA oxidation, nitrative stress, and advanced glycation end-products displayed more sustained reductions. These patterns indicate that the response was neither uniform across pathways nor uniformly durable over time. Dose-stratified analyses revealed little evidence of a consistent dose-response relationship. These findings are consistent with, but do not conclusively establish, a pharmacologically-driven antioxidant effect of NR, plausibly related to the known roles of NAD+ in supporting antioxidant defense systems. Indeed, independent preclinical studies, and at least one human study, have reported antioxidative properties of NR in various model systems, which may underie the health and therapeutic benefits observed with oral NR ^42–47^. Further, the antioxidative promoting effects of NR may at least partially explain the anti-inflammatory effects observed in previous clinical studies evaluating oral NR, as chronic oxidative stress is a known contributor to the development of low-grade persistent inflammation^48^.

The MeScreen assay provides a systems-level assessment of mitochondrial health and enables longitudinal evaluation of physiological responses to disease, lifestyle, or therapeutic interventions. A noteworthy finding was that the MitoATP/GlycoATP ratio increased relative to study baseline and remained elevated at Day 90. This shift was driven primarily by a reduction in GlycoATP, whereas MitoATP did not increase significantly. These results are suggestive of a change in the relative balance between oxidative and glycolytic pathway derived energy production. The concurrent reductions in basal ROS may reflect improved mitochondrial coupling efficiency, whereby enhanced NAD+ availability facilitates more complete electron transfer. Importantly, ROS levels did not increase despite enhanced mitochondrial activity, and the robust capacity for stress response was preserved, indicating that the metabolic shift did not compromise cellular redox balance. Overall, these findings are congruent with a preclinical rodent study demonstrating enhanced mitochondrial function and oxidative capacity upon oral NR administration, and partially align with oral NR human studies reporting alterations in substrate utilization and mitochondrial biogenesis^12 37,49^. In general, compromised oxidative energy production is a recognized feature of aging and insulin resistance, likely mediated in part by impaired mitochondrial content and function. These findings may therefore reflect improved mitochondrial homeostasis and metabolic flexibility ^50–52^. However, important to note is that oral NR’s effects on mitochondrial function parameters are not universally consistent. While mitochondrial biogenesis was demonstrated in Lapatto et al. following five months of NR67, other trials that have evaluated other indicators of mitochondrial biogenesis, such as PGC-1α protein, mtDNA copy number, citrate synthase activity, and OXPHOS complex content in human trials have had mixed results. For example, Dollerup et al.^53^ and Remie et al.^49^ found no improvement in mitochondrial respiration or content in human muscle after oral NR.

Dose-stratified analyses provided limited evidence of differential responses between the 50 mg and 100 mg groups. Although several outcomes demonstrated significant changes when analyzed collectively, within-dose analyses were generally less consistent. The observed loss of significance likely reflects reduced statistical power rather than an absence of dose-response effects, as the study was not powered to detect difference between doses. For mitochondrial outcomes, neither dose group displayed significant longitudinal changes across timepoints when analyzed separately. Urinary oxidative stress markers were broadly similar between dose groups. Collectively, though methodologically limited, the findings herein provide no clear support for a dose-dependent relationship within the ranges explored.

Sex-stratified analyses identified some differences in mitochondrial and oxidative stress outcomes. Within the mitochondrial assessment, male participants showed longitudinal changes in mitochondrial potential and stressed-minus-basal network measures. On the other hand, females demonstrated higher basal ROS at Day 90 and an increase in stressed ROS relative to baseline. With respect to urinary oxidative stress markers, female participants interestingly demonstrated a greater number of reductions in the measured parameters at Day 30 and/or Day 90 compared to baseline, while fewer statistically significant changes were evident among male participants. Collectively, sex-stratified analyses yielded mixed and directionally inconsistent findings, and should be considered exploratory.

The PRO findings provide preliminary, exploratory evidence of symptomatic benefits associated with NR injections across measures of fatigue, sleep, and QoL, though there was appreciable heterogeneity in responses. For instance, while sleep-related impairment was significantly reduced at Day 90 in the 100 mg group, no significant changes were evident in the 50 mg group, and sleep disturbance scores showed primarily directional changes without statistical significance. Global Mental Health scores improved significantly in the 50 mg group, while Global Physical Health scores did not significantly change. In contrast, fatigue scores were significantly lower by Day 90 in both groups. The interpretation of these findings is limited by the open-label design and a lack of a placebo control. It is well-established that subjective endpoints are susceptible to expectancy bias, and consequently, these findings should be considered exploratory signals at best that inform future placebo-controlled tests. Nevertheless, these findings are broadly aligned with emerging clinical evidence suggesting that NAD+ augmentation may influence symptom domains related to fatigue and sleep, although the existing evidence base for oral NR itself is mixed. In a 24-week, double-blind, placebo-controlled trial of oral NR in individuals with long-COVID, Wu et al., 2025 reported that 2000 mg/day of NR increased whole-blood NAD+ levels within 5 weeks; exploratory within-group analyses showed improvements in fatigue severity, sleep quality, mood, and executive functioning after 10 weeks of NR supplementation^54^. In the present study, improvements in fatigue and sleep emerged by Day 30 and were sustained through Day 90, suggesting that repeated parenteral NR administration may produce patient-reported benefits over a similar subchronic time frame.

Self-reported skin outcomes also represent an exploratory domain in which improvements were observed. The proportion of participants reporting facial skin improvement increased from 11.4% on Day 1 to 36.8% at Day 90. “Facial improvement noticed by others” and “body skin improvement” domains also increased progressively throughout the assessment timepoints. It should also be noted that a small proportion of participants reported unfavorable changes, including dryness, itchiness, or breakouts. Although NAD+ repletion in skin has been proposed to confer therapeutic and cosmetic benefits^55–57^, the findings of the present study should be interpreted as preliminary signals since they are reports of open-ended, subjective change assessments. Future studies should utilize validated, objective measures in blinded and controlled conditions to delineate the treatment effects of injectable NR on skin physiology.

From a translational standpoint, the present data strongly support the feasibility of injectable NR delivery. Unlike IV infusions, which require clinical supervision, SC administrations were successfully self-administered at home by participants three times weekly for 90 days, with low rates of injection-site related adverse findings, and no participant-withdrawals. This may be relevant given that oral NR bioavailability may be attenuated by gastrointestinal bioconversion; injections, on the other hand, may provide individuals with consistent, systemic exposure to intact NR from a route that bypasses first-pass metabolism. However, this does not necessarily mean that injected NR is definitively superior to oral or IV administration for clinical outcomes, and comparative bioavailability across oral, IV, IM, and SC routes remains to be formally established.

This study has several limitations that warrant consideration when interpreting these results. The open-label design and absence of a placebo control arm are particularly relevant for PROs, as subjective endpoints such as fatigue, sleep quality, pain, and QoL are susceptible to expectancy effects. Moreover, because participant eligibility required elevated baseline fatigue, the observed longitudinal improvements in PROs may reflect regression to the mean. Additionally, the present study’s small sample size limits statistical power for between-group comparisons and subgroup analyses. The extended, unsupervised at-home part also introduced potential variability in lifestyle (i.e., diet, exercise, concurrent supplement use, etc.) that was not independently verified beyond participant self-report. Of note, the at-home phase coincided with the U.S. winter holiday season, during which notable changes to lifestyle occur that coincide with untoward health changes^58,59^ Despite these limitations, the convergence of improvements across multiple independent endpoints—objective biomarkers (oxidative stress, mitochondrial function) and subjective outcomes (fatigue, sleep, pain, QoL)—strengthens the plausibility of a true biologic effect.

## 5. Conclusion

In the present study, 90 days of self-administered NR subcutaneously at 50 or 100 mg doses three times weekly were safe and well-tolerated in healthy adults presenting with elevated fatigue at baseline. There were no evident safety concerns across vital signs, clinical chemistry, hematology, or inflammatory markers. Whole-blood NAD+ concentrations remained relatively stable during the at-home phase of the study. Despite the absence of a measurable increase in whole blood NAD+, exploratory findings identified possible improvements in fatigue, sleep-related impairment, and quality of life perceptions. In parallel, a change in the MitoATP/GlycoATP ratio may signal a preferential shift toward oxidative phosphorylation-derived energy production. Additionally, there were reductions in the majority of oxidative stress biomarkers assessed. These findings, though exploratory, are directionally consistent with the known physiological roles of NAD+, and align with the oral NR literature. Therefore, the convergence of clinical and molecular endpoints provides preliminary evidence that injectable NR modulates NAD+-dependent cellular pathways with clinically relevant downstream effects. However, given the open-label design, modest sample size, and exploratory nature of the biological and functional endpoints, it bears emphasis that the longitudinal changes observed in the present study should be interpreted as hypothesis-generating, rather than estimates of treatment efficacy. Nonetheless, these findings warrant further investigation in randomized, placebo-controlled trials with larger sample sizes.

## Data Availability

All data produced in the present study are available upon reasonable request to the authors

## Author Contributions

AR– investigation, project administration, methodology, resources, software, supervision, visualization, writing – original draft, reviewing and editing;

YNE – conceptualization, funding acquisition, methodology, project administration, resources, supervision, writing – original draft, reviewing, and editing

FH – analysis, reviewing, and editing

JK – supervision, writing – original draft, reviewing, and editing

TA – investigation, writing – original draft, reviewing, and editing

AS – Funding acquisition, resources, writing – reviewing and editing

HC – Conceptualization, funding acquisition, project administration, resources, supervision, writing – review and editing

